# Comparative Effectiveness of the mRNA-1273 and BNT162b2 COVID-19 Vaccines Among Adults With Underlying Medical Conditions: A Systematic Literature Review and Pairwise Meta-Analysis Using GRADE

**DOI:** 10.1101/2024.09.13.24313632

**Authors:** Xuan Wang, Ankit Pahwa, Mary T. Bausch-Jurken, Anushri Chitkara, Pawana Sharma, Mia Malmenäs, Sonam Vats, Michael Gordon Whitfield, Kira Zhi Hua Lai, Priyadarsini Dasari, Ritu Gupta, Maria Nassim, Nicolas Van de Velde, Nathan Green, Ekkehard Beck

## Abstract

**Introduction:** This systematic literature review and pairwise meta-analysis evaluated the comparative effectiveness of mRNA-1273 versus BNT162b in patients with at least one underlying medical condition at high risk for severe COVID-19.

**Methods:** MEDLINE, Embase, and Cochrane databases were searched for relevant articles from January 1, 2019 to February 9, 2024. Studies reporting effectiveness data from at least two doses of mRNA-1273 and BNT162b2 vaccination in adults with medical conditions at high risk of developing severe COVID-19 according to the US Centers for Disease Control and Prevention were included. Outcomes of interest were SARS-CoV-2 infection (overall, symptomatic, and severe), hospitalization due to COVID-19, and death due to COVID-19. Risk ratios (RRs) were calculated with random effects models. Subgroup analyses by specific medical conditions, number of vaccinations, age, and SARS-CoV-2 variant were conducted. Heterogeneity between studies was estimated with chi-square testing. The certainty of evidence was assessed using the Grading of Recommendations, Assessments, Development, and Evaluations framework.

**Results:** Sixty-five observational studies capturing the original/ancestral-containing primary series to Omicron-containing bivalent original-BA4-5 vaccinations were included in the meta-analysis. mRNA-1273 was associated with significantly lower risk of SARS-CoV-2 infection (RR, 0.85 [95% CI, 0.79–0.92]; *I^2^*=92.5%), symptomatic SARS-CoV-2 infection (RR, 0.75 [95% CI, 0.65–0.86]; *I^2^*=62.3%), severe SARS-CoV-2 infection (RR, 0.83 [95% CI, 0.78–0.89]; *I^2^*=38.0%), hospitalization due to COVID-19 (RR, 0.88 [95% CI, 0.82–0.94]; *I^2^*=38.7%), and death due to COVID-19 (RR, 0.84 [95% CI, 0.76–0.93]; *I^2^*=1.3%) than BNT162b2. Findings were generally consistent across subgroups. Evidence certainty was low or very low because sufficiently powered randomized controlled trials are impractical in this heterogeneous population.

**Conclusion:** Meta-analysis of 65 observational studies showed that vaccination with mRNA-1273 was associated with a significantly lower risk of SARS-CoV-2 infection and COVID-19-related hospitalization and death than BNT162b2 in patients with medical conditions at high risk of severe COVID-19.

## INTRODUCTION

Although immunity to COVID-19 acquired through vaccination or natural SARS-CoV-2 infection has reduced rates of severe disease in the general population [1,2], the risk of COVID-19–associated morbidity and mortality in the post-pandemic setting remains high, particularly in people with specific medical conditions [3]. An estimated 1.7 billion people (22% of the global population) have at least one underlying medical condition, such as chronic lung diseases, cancer, chronic kidney disease, diabetes, heart conditions, or solid organ transplant, that increases their risk of developing severe COVID-19 [3,4]. Consistent with these risks, the World Health Organization (WHO) considers adults with these comorbidities to be a high priority group for COVID-19 vaccination [5].

The high efficacy and safety of the two mRNA vaccines developed against the ancestral SARS-CoV-2 strain, mRNA-1273 (Spikevax^®^; Moderna, Inc., Cambridge, MA, USA) [6] and BNT162b2 (Comirnaty^®^; Pfizer/BioNTech, New York, NY, USA/Mainz, Germany), [7] were demonstrated in phase 3 randomized controlled trials (RCTs) [8,9]. While immunocompromised individuals were excluded, the pivotal studies included participants with some high-risk comorbidities, and demonstrated high efficacy in these populations comparable to the healthy participants [8,9].

Although mRNA-1273 and BNT162b2 are both mRNA vaccines, their formulations differ in the amount of mRNA encoding the spike protein (mRNA-1273, 100/50 mcg; BNT162b2, 30 mcg) as well as in the lipid nanoparticle delivery systems [6,7,10,11]. In real-world studies differences in immunogenicity and effectiveness have been observed particularly in some high-risk populations, such as older and immunocompromised individuals [12–14]. Real-world effectiveness data are crucial to help inform vaccine selection and procurement decisions for COVID-19 national immunization programs because comparative high quality RCT evidence is lacking in the populations at greatest risk of severe COVID-19 disease.

Similar to seasonal influenza vaccination [15], COVID-19 vaccines are now updated periodically [16]; regulators and policy makers no longer require RCT evidence for approval of updated COVID-19 vaccines [17,18], and data from observational studies are increasingly used to support policy [19]. Given differences across vaccine formulations, the prohibitive number of people needed to enroll in potential RCTs designed for each high-risk group, and the variability in SARS-CoV-2 variants that emerge and their impact on the burden of disease, synthesis of effectiveness data using robust meta-analyses on the comparative performance of the two major COVID-19 vaccines provides a means to inform the choice of vaccine for future mRNA COVID-19 vaccination, particularly among vulnerable patient populations.

We therefore conducted a systematic literature review (SLR) and pairwise meta-analysis to compare the clinical effectiveness of mRNA-1273 versus BNT162b2 in adults with at least one underlying medical condition predisposing them to developing severe COVID-19. Our specific research question was “Following primary series vaccination and additional vaccine doses, is mRNA-1273 more effective than BNT162b2 at preventing SARS-CoV-2 infections and COVID-19–related hospitalizations and deaths in people with underlying medical conditions?” To evaluate the certainty of evidence generated from our meta-analysis, we employed the Grading of Recommendations, Assessments, Development, and Evaluations (GRADE) framework [20] used by national immunization technical advisory groups (NITAGs) to develop vaccine recommendations [21].

## METHODS

### Search strategy and study selection

This SLR and meta-analysis is registered in INPLASY (INPLASY202460065) and was conducted according to the Preferred Reporting Items for Systematic Reviews and Meta-Analyses 2020 framework [22]. Data are from previously conducted studies; no new studies with human participants or animals were performed.

Embase, MEDLINE, MEDLINE In-Process, e-pubs ahead of print, and Cochrane databases, including Cochrane Central Register of Controlled Trials and Cochrane Database of Systematic Reviews, were searched via the Ovid platform to retrieve studies published from January 1, 2019 to February 9, 2024. Search queries are provided in **Table S1**. To complement the main search, previous SLRs completed by our research group [12,13] as well as recently published SLRs were cross-checked for additional references [12,13,23–38].

The population, intervention, comparison, and outcomes used in the SLR are summarized in **Table S2.** We focused this SLR on adults with medical conditions known to increase the risk of developing severe COVID-19, a population considered to be a high priority group for COVID-19 vaccination by WHO and national public health institutes such as the US Centers for Disease Control and Prevention (CDC) and the European Centre for Disease Prevention and Control [3,5,39]. We defined the target population by selecting comorbidities from the list of medical conditions published by the CDC as those conferring elevated risk for developing severe COVID-19 [3]: adults with autoimmune disease, solid tumors, solid organ transplant, hematologic malignancies, chronic kidney disease with or without hemodialysis, type 1 or 2 diabetes, cardiovascular disease, cerebrovascular disease, chronic liver disease, neurologic disease, chronic respiratory disease, obesity, primary immunodeficiency syndrome or described as immunocompromised. Published clinical trials and observational studies reporting clinical effectiveness outcomes in adults ≥18 years of age with the previously specified underlying medical conditions were eligible for inclusion in the SLR.

Only studies reporting data from participants who received mRNA-1273 or BNT162b2, including variant-updated versions, were included. Homologous primary series with or without homologous or heterologous additional doses where the last dose was mRNA-1273 or BNT162b2 were allowed (**Table S2**). Studies reporting data from heterologous 2-dose vaccine regimens, comparing only mRNA-1273 or only BNT162b2 as the last dose of the series versus another COVID-19 vaccine as the last dose, or comparing mRNA-1273 or BNT162b2 primary series versus other COVID-19 vaccine primary series were excluded.

Clinical effectiveness outcomes of interest were those relevant to vaccine effectiveness (VE) against COVID-19 as follows: SARS-CoV-2 infection, symptomatic SARS-CoV-2 infection, severe SARS-CoV-2 infection, hospitalization due to COVID-19, and death due to COVID-19. SARS-CoV-2 infections were symptomatic or asymptomatic and confirmed by a positive laboratory test. Symptomatic infections were further defined as a positive test accompanied by COVID-19 symptoms, including but not limited to fever, cough, shortness of breath, sudden onset of anosmia or ageusia, and in some countries, runny nose. Severe SARS-CoV-2 infections were defined by individual studies; infections that resulted in hospitalization or death were also considered to be severe infections for this analysis. Hospitalization due to COVID-19 was defined as hospitalization associated with a positive SARS-CoV-2 test. Death due to COVID-19 was defined as death associated with a positive SARS-CoV-2 test and no other reported cause of death.

Additional rules used to determine study and data inclusion in the meta-analysis are provided in the **Appendix – Supplementary Methods**.

Two independent reviewers screened all identified articles using a 2-level approach. Discrepancies were resolved by consensus or a third reviewer. In level 1, titles and abstracts were screened against the inclusion criteria. In level 2, the full texts of articles that passed level 1 screening were retrieved and evaluated against selection criteria to determine final eligibility.

### Data extraction and quality assessment

Study design details, baseline patient and disease characteristics, vaccine intervention details, and effectiveness endpoints were extracted. Data were extracted for single, specific medical conditions if available; however, patients may have had other underlying comorbidities. Original study authors were contacted during data abstraction to provide clarity or additional data if needed.

Risk of bias was assessed in accordance with Cochrane review guidelines [40] using the Newcastle-Ottawa Scale [41] for observational studies. The certainty of evidence for meta-analysis results was evaluated based on GRADE [21].

### Statistical analysis

Consistent with previous comparative effectiveness meta-analyses in immunocompromised populations and older adults [12–14], we conducted a pairwise meta-analysis to compare vaccination with mRNA-1273 to vaccination with BNT162b2 in patients with underlying medical conditions. Only studies that reported the number of events and sample size, VE, or other effect measures per arm and subgroup were included in the meta-analysis. Risk ratios (RRs) were pooled across studies using random effects meta-analysis models. Individual studies were weighted using the inverse variance method [42]. A continuity correction of 0.5 was added to the number of events and sample size in both arms in studies where no events were observed in one or both arms [43]. A standard pairwise meta-analysis with RRs as the input comparing mRNA-1273 to BNT162b2 was conducted [42,44,45]. Where available, RRs were calculated from the number of events and sample size per arm, consistent with the methodology of other recently conducted meta-analyses for COVID-19 vaccines [28,34]. For studies that only reported VE, RR was estimated as 1−(VE/100) based on the reported definition of VE. Based on the assumption that all COVID-19 outcomes were rare [46,47], odds ratio (OR), hazard ratio (HR), and incidence rate ratio (IRR) were considered to be approximately equal to RR. The same approach was therefore taken for studies reporting OR, HR, or IRR; RR was estimated as (1–OR) × 100, (1–HR) × 100, and (1–IRR) × 100. If a study reported VE, OR, HR, or IRR both adjusted for covariates and unadjusted, adjusted data were preferentially used in the meta-analysis.

Because the target population consists of patients who may have multiple comorbidities and immunocompromising conditions, we primarily considered two populations: the broad at-risk population of adults with at least one medical condition as previously specified and the very high risk population of immunocompromised adults with medical conditions defined as clinically extremely vulnerable (CEV) groups 1 or 2 [48]. Although limited by feasibility, for comprehensiveness, we also conducted additional meta-analyses for specific groups and medical conditions (cancer, cardiovascular-metabolic-renal disease, treatment with immunosuppressive therapy) and individual comorbidities (autoimmune disease, cardiovascular disease, chronic kidney disease, chronic respiratory disease, diabetes, hematologic malignancy, solid organ transplant), which are presented in the **Appendix – Supplemental Methods** with the definitions of the medical conditions.

Similar to seasonal influenza vaccination, multiple mRNA COVID-19 vaccine formulations have been administered across pandemic and non-pandemic settings and COVID-19 vaccines are also expected to be periodically updated to match circulating variants [49]. In order to capture the variability of vaccine administration and updates over time, we performed subgroup analyses for the primary populations to evaluate the comparative effectiveness of mRNA-1273 versus BNT162b2 considering differences in dosing regimen (2 doses, ≥3 doses overall, ≥4 doses), receipt of homologous versus heterologous vaccine (applicable for ≥3 dose series), age group (18–65 years), and SARS-CoV-2 variant status (Delta, Omicron).

If the SARS-CoV-2 variant was not reported or specified directly, variants were assumed to be Delta from May 2021 (non-US studies) or June 2021 (US studies) through November 2021 and Omicron as of December 2021 based on the predominant variant circulating locally when data were collected [50–52].

If there were fewer than two studies for a given outcome or subgroup, the results of the corresponding analysis were not feasible.

Publication bias was assessed by visual examination of funnel plots and Egger’s regression test for asymmetry [53,54]. Heterogeneity across studies was evaluated using chi-square testing [55], with the percentage of variation across studies estimated using the *I^2^* statistic (scale, 0–100%; 0% indicates no evidence of heterogeneity). Results were summarized in forest plots showing effect estimates and their 95% CIs. Meta-analyses were conducted in R v4.3.1 using the packages meta [56] and metafor [57].

## RESULTS

### Search results and included studies

Of 3,814 articles screened, 68 were included in the SLR. Three studies were excluded from the meta-analysis because of insufficient data [58], mixed series VE [59], and unclear vaccine series with no clarification provided by authors [60], leaving 65 studies (all non-RCTs) included in the meta-analysis of adults with at least one underlying medical condition. Forty-five studies (all non-RCTs) of adults with CEV 1 or 2 conditions were included in the meta-analysis.

The characteristics of the 65 studies included in the meta-analysis are summarized in **Table 1**. Overall, our meta-analysis included over 4.0 million patients who received mRNA-1273 and over 5.1 million patients who received BNT162b2. Eleven studies were industry-sponsored [61–71]. More than half of the included studies were conducted in North America (United States, n=36 [62–64,66,67,70–100]; Canada, n=2 [101,102]; Mexico, n=1 [103]). Twenty-two studies involved European patients (Spain, n=9 [61,68,104–110]; Italy, n=5 [111–115]; Bulgaria, n=2 [116,117]; Austria, n=1 [118];Greece, n=1 [119]; Netherlands, n=1 [120]; Sweden, n=1 [121], Switzerland, n=1 [65]; multicountry, n=1 [122]). One study each was conducted in Qatar [69], Singapore [123], Taiwan [124], and in multiple countries [125]. Most studies involved patients with autoimmune disease (n=17) [63,65,74,76,80,85,87,91–93,95,97,104,112,115,120,123]; with mixed conditions (n=8 [67,71,76,86,90,101,118,122]), solid organ transplant (n=12 [64,72,81,88,99,100,105,106,111,119,121,124]), and chronic kidney disease (n=9 [62,67–69,73,89,96,102,125]) also common among included studies. Seven studies included populations with mixed cancer [64,70,75,78,83,84,94], with a further 8 studies including only patients with hematologic malignancies [64,70,76,98,107–110] and 5 studies including only patients with solid tumors [61,76,113,114,116]. Other medical conditions included were immunocompromised patients (n=5) [64,66,67,76,77], diabetes (n=5) [67,75,79,103,117], chronic respiratory disease (n=3) [67,75,116], cardiovascular disease (n=3) [67,75,116], obesity [103], primary immunodeficiency syndrome [64], and chronic liver disease (n=1) [82]. Most studies reported data from patients infected with the Delta variant (n=19), with studies reporting Omicron infection data also comprising a considerable proportion of the overall meta-analysis set (n=11). Risk of bias was not evaluated in 7 studies because full texts were not published [65,71,75,81,90,97,125]. Of the 58 remaining evaluable studies, 33 (57%) had no serious risk of bias and 24 (41%) had serious and 1 (2%) had very serious risk of bias (**Table S3**).

**Table 1.** Characteristics of included studies.

| Author, year | Study characteristics |  |  |  |  |  |  |  | Outcomes reported |  |  |  |  |
| --- | --- | --- | --- | --- | --- | --- | --- | --- | --- | --- | --- | --- | --- |
|  | Design | Country and data source | Medical condition | SARS-CoV-2 testing method | Variant | Vaccine doses | Study period | Vaccinated, n | SARS-CoV-2 Infection | Symptomatic SARS-CoV-2 infection | Severe SARS-CoV-2 infection | Hospitalization due to COVID-19 | Death due to COVID-19 |
| Alkadi, 2022 [69] | Test-negative case-control study | Qatar Hamad Medical Corporation | CKD with hemodialysis | PCR | NR | 2 doses (MM vs PP) | Dec 20, 2020 to Jan 3, 2022 | mRNA-1273: 69<br>BNT162b2: 622 | Y | N | N | N | N |
| Aslam, 2021 [72] | Retrospective cohort study | USA Transplant registry | Solid organ transplant | NS | Alpha<sup>a</sup> | 2 doses (MM vs PP) | Jan 2021 to Jun 2021 | mRNA-1273: 375<br>BNT162b2: 632 | Y | Y | Y | Y | Y |
| Bello-Chavolla, 2023 [103] | Retrospective cohort study | Mexico SISVER database | Diabetes, other (obesity) | RAT or RT-PCR | Mixed variants (B.1.1.5 19 and Delta) | 2 doses (MM vs PP) | Dec 24, 2020 to Sep 27, 2021 | **Diabetes**<br>mRNA-1273: 453<br>BNT162b2: 30,160<br>**Obesity**<br>mRNA-1273: 385<br>BNT162b2: 28,515 | Y | Y | Y | N | Y |
| Boekel, 2022 [120] | Pooled study from 2 prospective observational studies | Netherlands 2 large, ongoing prospective multicenter cohort studies | Autoimmune disease | RAT or PCR | Delta | 2 doses (MM vs PP) | Apr 26, 2020 to Mar 1, 2021; Feb 2, 2021 to Aug 1, 2021 | **With immunosuppressants**<br>mRNA-1273: 563<br>BNT162b2: 2,019<br>**Without immunosuppressants**<br>mRNA-1273: 203<br>BNT162b2: 598 | Y | N | Y | Y | Y |
| Bonazzetti, 2023 [111] | Prospective cohort study | Italy Medical records from RCCS Azienda Ospedaliero-Universitaria di Bologna | Solid organ transplant | RAT or PCR | Mixed variants (Alpha, Delta, and Omicron)<sup>a</sup> | 3 doses (MMM vs PPP; PPM vs PPP) | Feb 2021 to Jan 2022 (enrollment); follow-up until Apr 30, 2022 | mRNA-1273: 136<br>BNT162b2: 70 | Y | N | Y | Y | N |
| Butt, 2022 [73] | Test-negative case-control study | USA Department of Veterans Affairs COVID-19 Shared Data Resource | CKD with hemodialysis | Laboratory diagnosis | Mixed variants (Beta and Delta) | 2 doses (MM vs PP) | Jan 26, 2021 to Aug 31, 2021 | mRNA-1273: 1,174<br>BNT162b2: 1,526 | Y | N | N | N | N |
| Capuano, 2023 [112] | Retrospective cohort study | Italy MS Centers of the Raising | Autoimmune disease | RAT or PCR | Omicron<sup>a</sup> | 3 doses (MMM vs PPP; | Sep 2021 | mRNA-1273: 46 | N | N | Y | N | Y |
|  |  | Italian Researchers MS study group |  |  |  | PPM vs MMP) | to Jul 2022 | BNT162b2: 244 |  |  |  |  |  |
| Chen, 2023 [124] | Retrospective cohort study | Taiwan Taiwan Centers for Disease Control | Solid organ transplant | RAT or PCR | Omicron | 2 doses (MM vs PP)<br>3 doses (MMM vs PPP; PPM, AAM, XXM vs MMP, AAP, XXP)<br>4 doses (MMM M vs PPPP; MMPM, MMXM , PPMM, PPPM, AMMM , AAMM, AAPM, AAXM, XXMM vs MMMP, AAMP) | Apr to Aug 2022 | **2 doses**<br>mRNA-1273: 24<br>BNT162b2: 19<br>**3 doses**<br>mRNA-1273: 219<br>BNT162b2: 91<br>**4 doses**<br>mRNA-1273: 61<br>BNT162b2: 11 | Y | N | N | N | N |
| Cona, 2023 [113] | Prospective cohort study | Italy Patient questionnaire from Luigi Sacco Hospital | Solid tumor | RAT or PCR | Alpha<sup>a</sup> | 2 doses (MM vs PP) | Jan to Jun 2021 | mRNA-1273: 55<br>BNT162b2: 140 | Y | Y | N | N | N |
| Cook, 2023 [74] | Retrospective cohort study | USA<br>Electronic health records from Mass General Brigham | Autoimmune disease | RAT or PCR | Omicron and pre-Omicron (Alpha and Delta) | 2 doses (MM vs PP)<br>2 or 3 doses (MM or MMM vs PP or PPP) | Dec 27, 2020 to May 15, 2021; end of follow-up Feb 22, 2022 | mRNA-1273: 4,322<br>BNT162b2: 5,516 | Y | N | N | N | N |
| Dimitrov, 2023 [116] | Retrospective cohort study | Bulgaria<br>Bulgarian Ministry of Health United Information Portal | Cardiovascular disease, chronic respiratory condition, solid tumor | RT-PCR and RAT | Mixed variants | 2 doses (MM vs PP)<br>3 doses (AAM vs AAP; JM vs JP) | Mar 2020 to Apr 2022 | **2 doses**<br>mRNA-1273: 914<br>BNT162b2: 8,033<br>**3 doses**<br>mRNA-1273: 96<br>BNT162b2: 900 | N | N | Y | N | Y |
| Drawz, 2022 [75] | Test-negative case-control study | USA<br>Minnesota Electronic Health Record Consortium | Cardiovascular disease (including hypertension), chronic respiratory condition, diabetes, mixed cancer | PCR | Delta<sup>a</sup> | 2 doses (MM vs PP)<br>3 doses (MMM vs PPP) | Aug 29, 2021 to Nov 27, 2021 | **2 doses**<br>mRNA-1273: 1,066,645<br>BNT162b2: 1,732,112<br>**3 doses**<br>mRNA-1273: 609,153<br>BNT162b2: 395,634 | Y | N | Y | Y | N |
| Egri, 2023 [104] | Prospective cohort study | Spain<br>Department of Autoimmune Diseases,<br>Hospital Clinic Barcelona | Autoimmune disease | RAT or PCR | Mixed variants (Delta and Omicron)<sup>a</sup> | 3 doses (MMM vs PPP) | May 2021 to Jan 2022 | mRNA-1273: 7<br>BNT162b2: 2 | Y | Y | N | N | N |
| Embi, 2021 [76] | Test-negative case-control study | USA<br>VISION Network | Autoimmune disease, hematologic malignancy, immunocompromised, mixed conditions, solid tumor | PCR | Delta | 2 doses (MM vs PP) | Jan 17 to Sep 5, 2021 | mRNA-1273: 4,337<br>BNT162b2: 6,227 | Y | N | Y | Y | N |
| Embi, 2023 [77] | Test-negative case-control study | USA<br>VISION Network | Immunocompromised | RT-PCR | Delta | 2 doses (MM vs PP)<br>3 doses (MMM vs PPP) | Aug 26 to Dec 25, 2021 | **2 doses**<br>mRNA-1273: 1,698<br>BNT162b2: 2,509<br>**3 doses**<br>mRNA-1273: 452<br>BNT162b2: 919 | Y | N | Y | Y | N |
| Figueiredo, 2021 [78] | Prospective cohort study | USA<br>SeroNet CORALE study | Mixed cancer | SARS-CoV-2 IgG assays | Mixed variants (Alpha and Delta)<sup>a</sup> | 2 doses (MM vs PP) | Dec 22, 2020 to Aug 30, 2021 | mRNA-1273: 128<br>BNT162b2: 163 | Y | N | Y | Y | N |
| Fu, 2023 [79] | Retrospective cohort study | USA<br>National | Diabetes | PCR | Pre-Delta, Delta, Omicron | 2 doses (MM vs PP) | Dec 11, 2020 to Jun | **2 doses**<br>mRNA-1273: 55,234 | Y | N | Y | N | Y |
|  |  | COVID Cohort Collaborative |  |  | , and mixed variants | 3 doses (MMM vs PPP; PPM vs MMP; JM vs JP) | 30, 2022 | BNT162b2: 86,447<br>**3 doses**<br>mRNA-1273: 46,065<br>BNT162b2: 48,528 |  |  |  |  |  |
| Grewal, 2022 [101] | Test-negative case-control study | Canada Provincial databases | Mixed conditions | RT-PCR | Omicron | 3 doses (MMM vs PPP) | Dec 30, 2021 to Apr 27, 2022 | mRNA-1273: 57,604<sup>b</sup><br>BNT162b2: 48,706<sup>b</sup> | Y | Y | Y | N | N |
| Haarhaus, 2023 [125] | Retrospective cohort study | 22 countries in Europe, Asia, Africa, and South America Multinational dialysis provider network | CKD with hemodialysis | PCR | Mixed variants (Alpha and Delta)<sup>a</sup> | 2 doses (MM vs PP) | Jan 31 to Jul 15, 2021 | mRNA-1273: 1,521<br>BNT162b2: 13,116 | Y | N | N | N | N |
| Heinzl, 2022 [118] | Retrospective cohort study | Austria Electronic health records from Department of Internal Medicine at Saint John of God Hospital Linz | Mixed conditions | PCR | Delta | 2 doses (MM vs PP) | Aug to Dec 2021 | mRNA-1273: 4<br>BNT162b2: 61 | N | N | Y | N | Y |
| Hernandez, 2022 [61] | Prospective cohort study | Spain Medical records | Solid tumor | PCR | Mixed variants (Alpha, Delta, and | 2 doses (MM vs PP) | NR; censor data Jun 2, 2022 | **2 doses**<br>mRNA-1273: 119 | N | N | Y | N | Y |
|  |  |  |  |  | Omicron)<sup>a</sup> | 3 doses (MMM vs PPP) |  | BNT162b2: 2<br>**3 doses**<br>mRNA-1273: 93<br>BNT162b2: 1 |  |  |  |  |  |
| Holroyd, 2022 [80] | Retrospective cohort study | USA Brigham MS Center CLIMB repository | Autoimmune disease | PCR, anti-Spike antibody testing (Roche Elecsys) | Delta<sup>a</sup> | 2 doses (MM vs PP) | Jun to Dec 2021 | mRNA-1273: 110<br>BNT162b2: 133 | Y | N | Y | Y | Y |
| Joerns, 2022 [81] | Retrospective cohort study | USA University of Texas Southwestern Medical Center | Solid organ transplant | NR | Mixed variants<sup>a</sup> | 2 doses (MM vs PP) | Mar 1, 2020 to Sep 24, 2021 | mRNA-1273: 18<br>BNT162b2: 33 | Y | Y | N | N | N |
| John, 2022 [82] | Retrospective cohort study | USA Veterans Outcomes and Costs Associated with Liver Disease cohort | Chronic liver disease | PCR | Mixed variants (Delta and Omicron) | 2 doses (MM vs PP)<br>3 doses (MMM vs PPP) | Dec 18, 2020 to Feb 11, 2022 | **2 doses**<br>mRNA-1273: 6,875<br>BNT162b2: 6,166<br>**3 doses**<br>mRNA-1273: 6,229<br>BNT162b2: 6,812 | N | N | Y | N | Y |
| Kelly, 2022 [84] | Retrospective cohort study | USA Veterans Health Administration | Mixed cancer | Laboratory diagnosis | Mixed variants (Delta and Omicron) | 3 doses (MMM vs PPP) | Jul 1, 2021 to Apr 29, 2022; follow-up until May 30, 2022 | mRNA-1273: 79,517<br>BNT162b2: 67,780 | Y | Y | Y | Y | N |
| Kelly, 2023 [83] | Retrospective cohort study | USA Veterans Health Administration COVID-19 Shared Data Resource | Mixed cancer | Laboratory diagnosis | Delta, Omicron, and mixed variants (Delta and Omicron) | 3 doses (MMM vs PPP) | Jul 1, 2021 to Apr 9, 2022; follow-up until May 30, 2022 | mRNA-1273: 101,749<br>BNT162b2: 87,308 | Y | Y | Y | N | N |
| Khan, 2021 [85] | Retrospective cohort study | USA Veterans Health Administration | Autoimmune disease | PCR, antibody testing, or physician note | Alpha<sup>a</sup> | 2 doses (MM vs PP) | Dec 18, 2020 to Apr 20, 2021 | mRNA-1273: 3,380<br>BNT162b2: 2,873 | Y | N | Y | N | N |
| Kissling, 2022 [122] | Test-negative case-control study | Europe (Croatia, France, Ireland, the Netherlands, Portugal, Romania, Spain [4 regions], England, | Mixed conditions | RT-PCR, antigen testing | Delta | 2 doses (MM vs PP) | Jul to Aug 2021 | mRNA-1273: 172<br>BNT162b2: 1,016 | Y | Y | N | N | N |
|  |  | Scotland)<br>Medical<br>records,<br>questionnaires,<br>vaccine registry |  |  |  |  |  |  |  |  |  |  |  |
| Kopel, 2024<br>[67] | Retrospective<br>cohort study | USA<br>Veradigm<br>Network EHR<br>dataset and<br>Komodo<br>dataset | Cardiovascul<br>ar disease,<br>chronic<br>kidney<br>disease with<br>and without<br>hemodialysis,<br>chronic<br>respiratory<br>condition,<br>diabetes,<br>immunocom<br>promised,<br>mixed<br>conditions | HCP<br>confirm<br>ed<br>diagnos<br>is codes | Omicron | ≥3 doses<br>(X<sup>c</sup>M vs<br>X<sup>c</sup>P) | Sep 8,<br>2022<br>to May<br>31,<br>2023 | mRNA-1273:<br>757,572<br>BNT162b2:<br>1,204,975 | Y | N | Y | Y | N |
| Kshirsagar,<br>2022 [86] | Retrospective<br>cohort study | USA<br>medical claims<br>data | Mixed<br>conditions | Laborat<br>ory<br>diagnos<br>is | Mixed<br>variants<br>(Alpha<br>and<br>Delta)<sup>a</sup> | 2 doses<br>(MM vs<br>PP) | Mar 10<br>to Oct<br>14,<br>2021 | mRNA-1273: 5,480<br>BNT162b2:<br>11,339 | N | N | Y | Y | Y |
| Liew, 2022 [87] | Retrospective<br>cohort study | USA<br>COVID-19<br>Global<br>Rheumatology<br>Alliance | Autoimmune<br>disease | PCR,<br>antigen<br>or<br>antibod<br>y test | Mixed<br>variants<br>(Alpha<br>and<br>Delta)<sup>a</sup> | 2 doses<br>(MM vs<br>PP) | Jan 5<br>to Sep<br>30,<br>2021 | mRNA-1273: 21<br>BNT162b2:<br>45 | N | N | Y | Y | Y |
| Magnusson,<br>2021 [121] | Retrospective<br>cohort study | Sweden<br>Referring<br>hospitals in<br>Sweden | Solid organ<br>transplant | PCR,<br>antigen<br>test | Mixed<br>variants<sup>a</sup> | 2 doses<br>(MM vs<br>PP) | Feb 1,<br>2020<br>to Apr<br>30,<br>2021 | mRNA-1273: 3<br>BNT162b2:<br>4 | Y | Y | Y | Y | N |
| Malinis, 2021 [88] | Retrospective cohort study | USA<br>Yale New Haven Hospital | Solid organ transplant | NR | Mixed variants (Alpha and Delta)<sup>a</sup> | 2 doses (MM vs PP) | Start date NR; end date as of May 18, 2021 | mRNA-1273: 167<br>BNT162b2: 288 | Y | N | Y | N | Y |
| Manley, 2023 [62] | Retrospective cohort study | USA<br>EHR | CKD with hemodialysis | RT-PCR | Delta, pre-Delta, and mixed variants (Delta and pre-Delta) | 2 doses (MM vs PP) | Feb 1 to Dec 18, 2021 | mRNA-1273: 6,853<br>BNT162b2: 5,132 | Y | N | Y | Y | Y |
| Marinaki, 2022 [119] | Prospective cohort study | Greece<br>Hospital records from Laiko Hospital Athens | Solid organ transplant | PCR | Mixed variants (Delta and Omicron)<sup>a</sup> | 2 doses (MM vs PP) | NR | mRNA-1273: 147<br>BNT162b2: 302 | Y | N | Y | Y | N |
| Mazuecos, 2022 [105] | Retrospective cohort study | Spain<br>Spanish Society of Nephrology COVID-19 Registry | Solid organ transplant | NR | Delta | 2 doses (MM vs PP) | Apr 1 to Oct 2, 2021 | mRNA-1273: 213<br>BNT162b2: 121 | N | N | Y | Y | Y |
| Miao, 2023 [89] | Retrospective cohort study | USA<br>Mayo Health Clinic in the Midwest | CKD with hemodialysis | Laboratory diagnosis | Mixed variants (Alpha, Delta, and Omicron) | 2 doses (MM vs PP) | Apr 1, 2020 to Oct 31, 2022 | mRNA-1273: 53<br>BNT162b2: 114 | N | N | Y | Y | Y |
| Motwani, 2023 [63] | Prospective cohort study | USA Data source NR | Autoimmune disease | PCR | Mixed variants (Delta and Omicron)<sup>a</sup> | 2 doses (MM vs PP) | >12 months | mRNA-1273: 684<br>BNT162b2: 1,170 | Y | N | Y | Y | N |
| Mues, 2022 [64] | Retrospective cohort study | USA HealthVerity database | Hematologic malignancy, immunocompromised, mixed cancer, other (primary immunodeficiency syndrome), solid organ transplant | PCR or antigen test | Delta | 2 doses (MM vs PP) | Dec 11, 2020 to Oct 12, 2021 | mRNA-1273: 57,700<br>BNT162b2: 66,757 | Y | N | Y | Y | N |
| Nguyen, 2023 [71] | Retrospective cohort study | USA EHR linked to pharmacy and medical claims | Mixed conditions | NR | Mixed variants (Delta and Omicron)<sup>a</sup> | 2 doses (MM vs PP)<br>3 doses (MMM vs PPP) | Primary series: Feb 1 to Oct 18, 2021<br>Booster: Oct 19, 2021 to Jan 31, 2022 | **2 doses**<br>mRNA-1273: 855,458<br>BNT162b2: 842,335<br>**3 doses**<br>mRNA-1273: 208,725<br>BNT162b2: 214,564 | Y | N | Y | Y | N |
| Niu, 2022 [90] | Retrospective cohort study | USA Memorial Healthcare System, | Mixed conditions | NR | Delta<sup>a</sup> | 2 doses (MM vs PP) | Jun 1 to Sep 20, 2021 | mRNA-1273: 37<br>BNT162b2: 225 | N | N | Y | N | Y |
|  |  | Hollywood, Florida |  |  |  |  |  |  |  |  |  |  |  |
| Odriozola, 2022 [106] | Retrospective cohort study | Spain<br>Medical records and patient surveys from Marqués de Valdecilla University Hospital and San Pedro Hospital | Solid organ transplant | RAT confirmed by RT-PCR or antigen test | Omicron | 3 doses (MMM vs MMP) | Late Nov 2021 to Feb 23, 2022 | mRNA-1273: 23<br>BNT162b2: 6 | N | N | Y | Y | Y |
| Parsons, 2023 [91] | Prospective cohort study | USA<br>Beth Israel Deaconess Medical Center lupus cohort | Autoimmune disease | PCR or anti-nucleocapsid IgG assay | Mixed variants (Delta and Omicron)<sup>a</sup> | 2 doses (MM vs PP)<br>3 doses (MMM vs PPP) | Dec 2020 to Mar 2022 | **2 doses**<br>mRNA-1273/BNT162b2: 16<sup>d</sup><br>**3 doses**<br>mRNA-1273: 27<br>BNT162b2: 35 | Y | N | N | N | N |
| Patel, 2023 [92] | Retrospective cohort study | USA<br>EHR from Mass General Brigham | Autoimmune disease | PCR or antigen test | Mixed variants (Delta and Omicron)<sup>a</sup> | 2 doses (MM vs PP) | Dec 11, 2020 to Nov 15, 2021 | mRNA-1273: 4,588<br>BNT162b2: 6,080 | Y | N | N | N | N |
| Pinana, 2022 [107] | Prospective cohort study | Spain<br>GETH-TC registry | Hematologic malignancy | PCR or antigen test | Delta<sup>a</sup> | 2 doses (MM vs PP) | Dec 2020 to early Dec 2021 | mRNA-1273: 983<br>BNT162b2: 362 | Y | N | N | N | N |
| Pinana, 2023a [109] | Prospective cohort study | Spain<br>GETH-TC registry | Hematologic malignancy | PCR or antigen test | Mixed variants (Alpha or Beta, Delta, and Omicron ) | 2 doses (MM vs PP)<br>3 doses (MMM vs PPP) | Primary: Dec 30, 2020 to Jun 30, 2021<br>Booster: until Jul 28, 2022 | **2 doses**<br>mRNA-1273: 1,086<br>BNT162b2: 410<br>**3 doses**<br>mRNA-1273: 910<br>BNT162b2: 361 | Y | N | N | N | N |
| Pinana, 2023b [108] | Prospective cohort study | Spain<br>GETH-TC registry | Hematologic malignancy | PCR or antigen test | Mixed variants (Alpha or Beta, Delta, and Omicron ) | 2 doses (MM vs PP)<br>3 doses (MMM vs PPP) | Primary: Dec 30, 2020 to Jun 30, 2021<br>Booster: until Jul 28, 2022 | **2 doses**<br>mRNA-1273: 456<br>BNT162b2: 97<br>**3 doses**<br>mRNA-1273: 97<br>BNT162b2: 382 | Y | N | N | N | N |
| Pino, 2022 [114] | Retrospective cohort study | Italy<br>Medical Oncology Unit in Florence at Santa Maria Annunziata, Serristori and Borgo San Lorenzo Hospitals | Mixed cancer | NR | Alpha<sup>a</sup> | 2 doses (MM vs PP) | Mar 26 to Apr 4, 2021 | mRNA-1273: 527<br>BNT162b2: 96 | Y | N | N | N | N |
| Quiroga, 2022 [68] | Prospective cohort study | Spain Hospital | CKD with hemodialysis | PCR or antigen test | NR | 3 doses (XXM vs XXP) | NR | mRNA-1273: 481<br>BNT162b2: 230 | Y | N | N | N | N |
| Radcliffe, 2022 [100] | Retrospective cohort study | USA Medical chart records from transplant center | Solid organ transplant | PCR | NR | 3 doses (MMM vs PPP)<br>4 doses (MMM M vs PPPP) | NR | **3 doses**<br>mRNA-1273: 142<br>BNT162b2: 282<br>**4 doses**<br>mRNA-1273: 1<br>BNT162b2: 2 | Y | N | N | N | N |
| Raptis, 2023 [65] | Prospective cohort study | Switzerland SCQM registry | Autoimmune disease | Patient-reported PCR or antigen test | Mixed variants (Delta and Omicron)<sup>a</sup> | 3 doses (MMM or RRM vs PPP or RRP) | Dec 8, 2021 to Nov 24, 2022 | mRNA-1273: 218<br>BNT162b2: 227 | Y | N | N | N | N |
| Risk, 2022 [93] | Retrospective cohort study | USA Michigan Medicine healthcare system | Autoimmune disease | PCR | Omicron | 2 doses (MM vs PP)<br>3 doses (MMM vs PPP) | Dec 16, 2021 to Mar 4, 2022 | mRNA-1273: 4,863<sup>d</sup><br>BNT162b2: 4,863<sup>d</sup> | Y | N | Y | Y | N |
| Rodriguez-Mora, 2023 [110] | Prospective cohort study | Spain EHR from patients with CML at Hospital Universitario Ramon y Cajal | Hematologic malignancy | Antigen test | Mixed variants (Alpha, Delta, and Omicron)<sup>a</sup> | 2 doses (MM vs PP) | Jan 2021 to 17 months after second dose | mRNA-1273: 16<br>BNT162b2: 9 | Y | N | N | N | N |
| Rooney, 2022 [94] | Retrospective cohort study | USA University of Kansas | Mixed cancer | PCR | Delta | 2 doses (MM vs PP) | Feb to Oct 2021 | mRNA-1273: 2,993<br>BNT162b2: 6,423 | Y | N | N | N | N |
| Shen, 2022 [95] | Retrospective cohort study | USA Michigan Medicine healthcare system | Autoimmune disease | Laboratory test | Delta | 2 doses (MM vs PP) | Jan 1 to Dec 7, 2021 | mRNA-1273: 2,046<br>BNT162b2: 2,064 | Y | N | Y | Y | N |
| Sibbel, 2022 [96] | Retrospective cohort study | USA Dialysis organization | CKD with hemodialysis | PCR | Alpha<sup>a</sup> | 2 doses (MM vs PP) | Jan 15 to Apr 30, 2021 | mRNA-1273: 19,900<br>BNT162b2: 10,140 | Y | N | Y | Y | Y |
| Singh, 2021 [97] | Retrospective cohort study | USA National COVID Cohort Collective | Autoimmune disease | NR | Mixed variants<sup>a</sup> | 2 doses (MM vs PP) | Jan 2020 to Sep 23, 2021 | mRNA-1273: 11,300<br>BNT162b2: 30,670 | Y | N | N | N | N |
| Song, 2022 [70] | Retrospective cohort study | USA National COVID Cohort Collective | <u>Hematologic malignancy</u>, <u>mixed cancer</u> | RT-PCR | Mixed variants (Alpha and Delta)<sup>a</sup> | 2 doses (MM vs PP) | Dec 1, 2020 to May 31, 2021 | mRNA-1273: 15,895<br>BNT162b2: 43,259 | Y | N | Y | N | N |
| Sormani, 2022 [115] | Prospective cohort study | Italy 35 Italian multiple sclerosis centers | Autoimmune disease | PCR | Delta, Omicron, and mixed variants (Delta and Omicron) | 2 doses (MM vs PP) | Mar 4 to Dec 15, 2021 | mRNA-1273: 314<br>BNT162b2: 1,391 | Y | N | N | N | N |
| Sun, 2024 [66] | Retrospective cohort study | USA HealthVerity database | Immunocompromised | NR | Mixed variants<sup>a</sup> | 3 doses (RRM vs RRP) | Dec 11, 2021 to Aug 31, 2022 | mRNA-1273: 52,943<br>BNT162b2: 60,084 | Y | N | Y | Y | N |
| Valkov, 2023 [117] | Retrospective cohort study | Bulgaria Bulgarian Ministry of Health United Information Portal | Diabetes | RT-PCR, antigen test | Mixed variants<sup>a</sup> | 2 doses (MM vs PP)<br>3 doses (JM vs JP; AAM vs AAP) | Mar 2020 to Jun 2022 | **2 doses**<br>mRNA-1273: 73<br>BNT162b2: 553<br>**3 doses**<br>mRNA-1273: 6<br>BNT162b2: 55 | N | N | Y | N | Y |
| Wang, 2022 [98] | Retrospective cohort study | USA EHR | Hematologic malignancy | NR | Mixed variants (Alpha and Delta)<sup>a</sup> | 2 doses (MM vs PP) | Dec 2020 to Oct 2021 | mRNA-1273: 1,239<br>BNT162b2: 4,658 | Y | N | Y | Y | N |
| Wing, 2023 [102] | Retrospective cohort study | Canada Provincial health administrative databases | CKD with hemodialysis | RT-PCR | Omicron | 3 doses (MMM vs PPP) | Dec 1, 2021 to Feb 28, 2022 | mRNA-1273: 1,662<br>BNT162b2: 4,761 | Y | N | N | N | N |
| Yeo, 2022 [123] | Prospective cohort study | Singapore National Neuroscience Institute and National University Hospital | Autoimmune disease | PCR, RAT | Delta<sup>a</sup> | 2 doses (MM vs PP)<br>3 doses (MMM vs PPP) | Aug 1, 2021 to Dec 31, 2021 | **2 doses**<br>mRNA-1273: 38<br>BNT162b2: 327<br>**3 doses** mRNA-1273:31<br>BNT162b2:141 | Y | Y | Y | Y | N |
| Yetmar, 2022 [99] | Retrospective cohort study | USA NR | Solid organ transplant | PCR, antigen test | Delta | 2 doses (MM vs PP) | Aug to Sep 2021 | mRNA-1273: 12<br>BNT162b2: 22 | N | N | Y | N | Y |

<sup>a</sup> Assumed based on study period.
<sup>b</sup> Number of vaccinated participants included in the infection analysis. For the symptomatic infection analysis, n=1,831 (mRNA-1273) and n=2,139 (BNT162b2; for the severe infection analysis, n=1,518 (mRNA-1273) and n=1,638 (BNT162b2).
<sup>c</sup> Undefined number of doses of any vaccine prior to the Omicron-containing bivalent original-BA4-5 booster dose.
<sup>d</sup> Overall number of vaccinated participants. No separate numbers per vaccine arm were provided.
A, dose of AstraZeneca ChAdOx1 COVID-19 vaccine; CKD, chronic kidney disease; CML, chronic myeloid leukemia; IgG, immunoglobulin G; J, dose of Johnson & Johnson Ad26.COV2.S COVID-19 vaccine; EHR, electronic health record; M, dose of Moderna mRNA-1273 COVID-19 vaccine; MS, multiple sclerosis; NR, not reported; NS, not specified; PCR, polymerase chain reaction; P, dose of Pfizer/BioNTech BNT162b2 COVID-19 vaccine; R, dose of any mRNA COVID-19 vaccine; RAT, rapid antigen test; RT-PCR, reverse transcription polymerase chain reaction; X, dose of unknown COVID-19 vaccine.

Meta-analysis results of the comparative effectiveness of mRNA-1273 and BNT162b2 in the base-case are shown in **Figures 2–5** and in the subgroups of groups of medical conditions and individual comorbidities in **Figures S1–S10**.

**Fig. 1.**
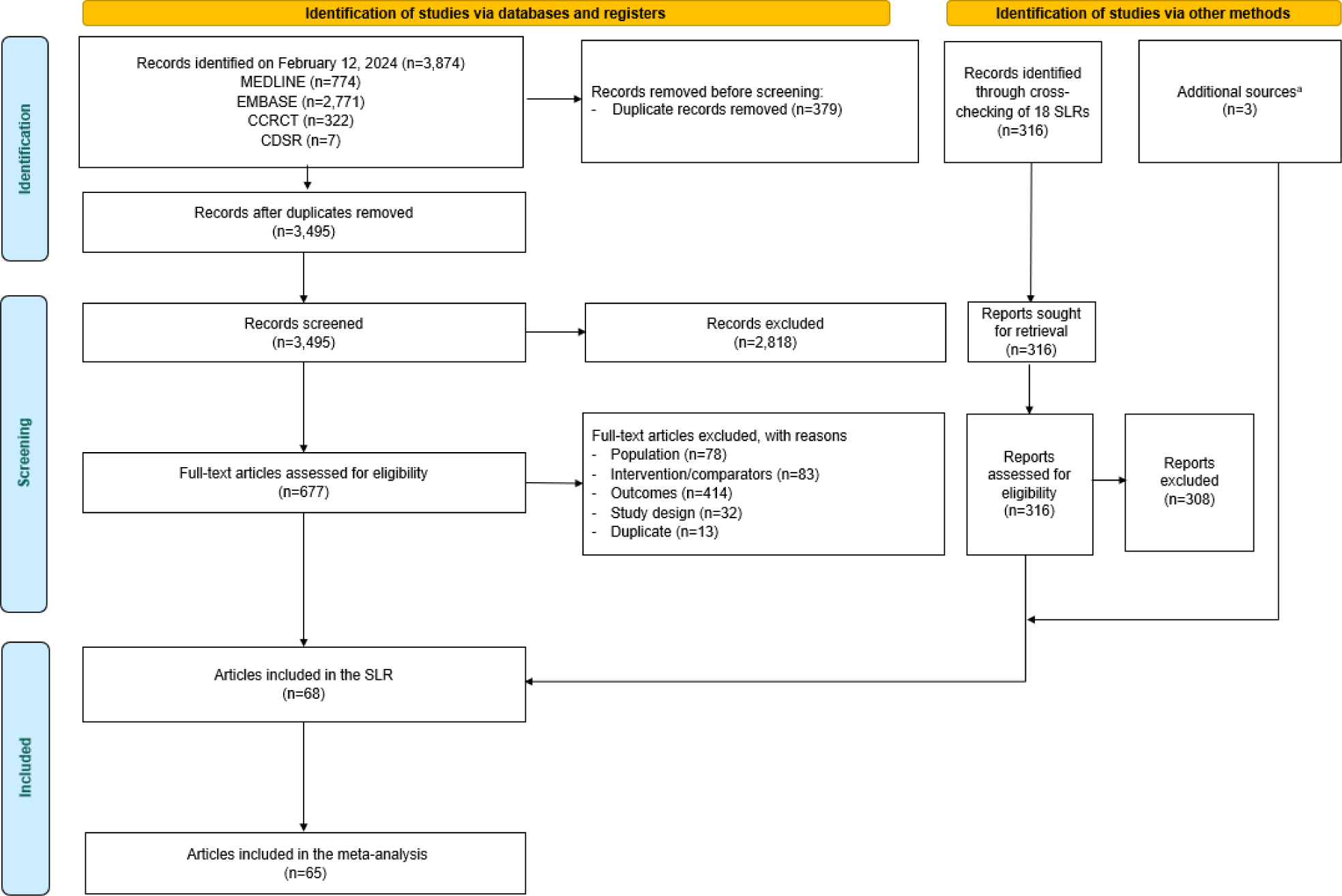
PRISMA flow diagram. ^a^ Includes 2 preprints and 1 poster. CCRT, Cochrane Central Register of Controlled Trials; CDSR, Cochrane Database of Systematic Reviews; PRISMA, Preferred Reporting Items for Systematic Reviews and Meta-Analyses; SLR, systematic literature review.

**Fig. 2.**
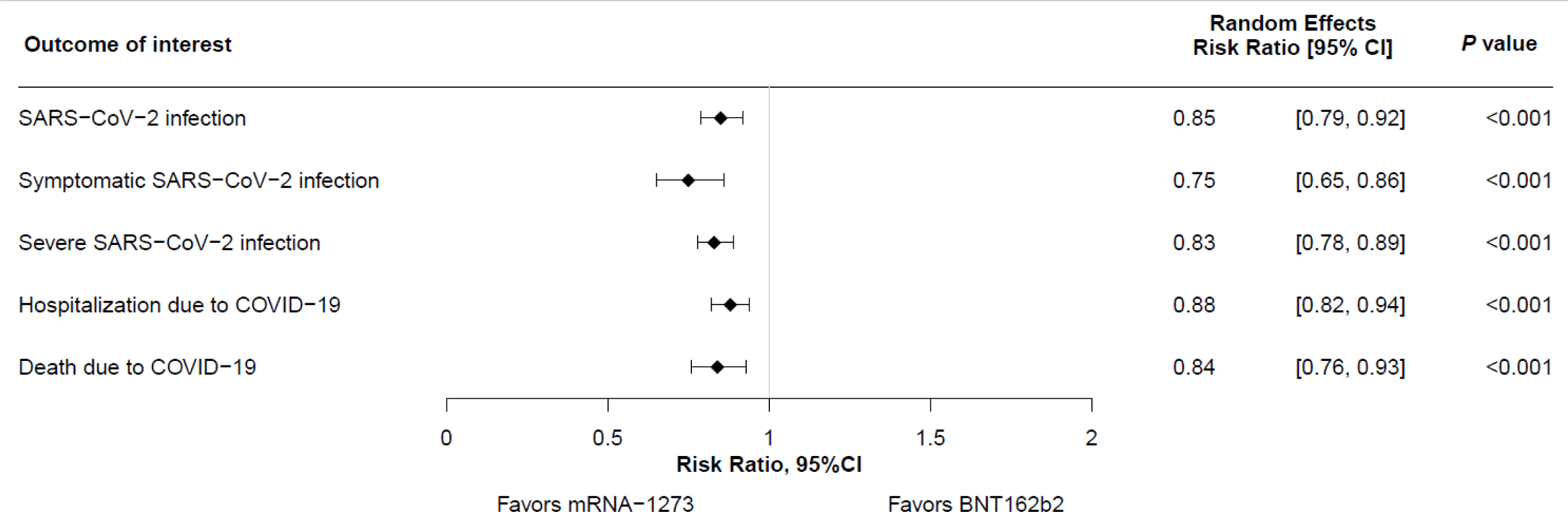
Summary of meta-analysis results on clinical effectiveness outcomes of the mRNA-1273 versus BNT162b2 COVID-19 vaccines in the overall population of adults with at least one of the following underlying medical conditions: autoimmune disease, solid tumor, solid organ transplant, hematologic malignancy, chronic kidney disease with and without hemodialysis, type 1 and 2 diabetes, cardiovascular disease, cerebrovascular disease, chronic liver condition, neurologic condition, chronic respiratory condition, obesity.

There was no suspected publication bias for any outcomes in the base-case populations, except for possible publication bias for symptomatic SARS-CoV-2 infection in adults with CEV 1 or 2 conditions (Egger’s regression test, *P* < 0.05).

### SARS-CoV-2 infection

Meta-analysis of 52 studies reporting the SARS-CoV-2 infection outcome found that vaccination with mRNA-1273 was associated with a significantly lower risk of SARS-CoV-2 infection compared with vaccination with BNT162b2 in adults with at least one underlying medical condition (RR, 0.85 [95% CI, 0.79–0.92]; **Figure 2** and **Figure 3a**). There was considerable heterogeneity between the studies (*I^2^*=92.5%).

**Fig. 3.**
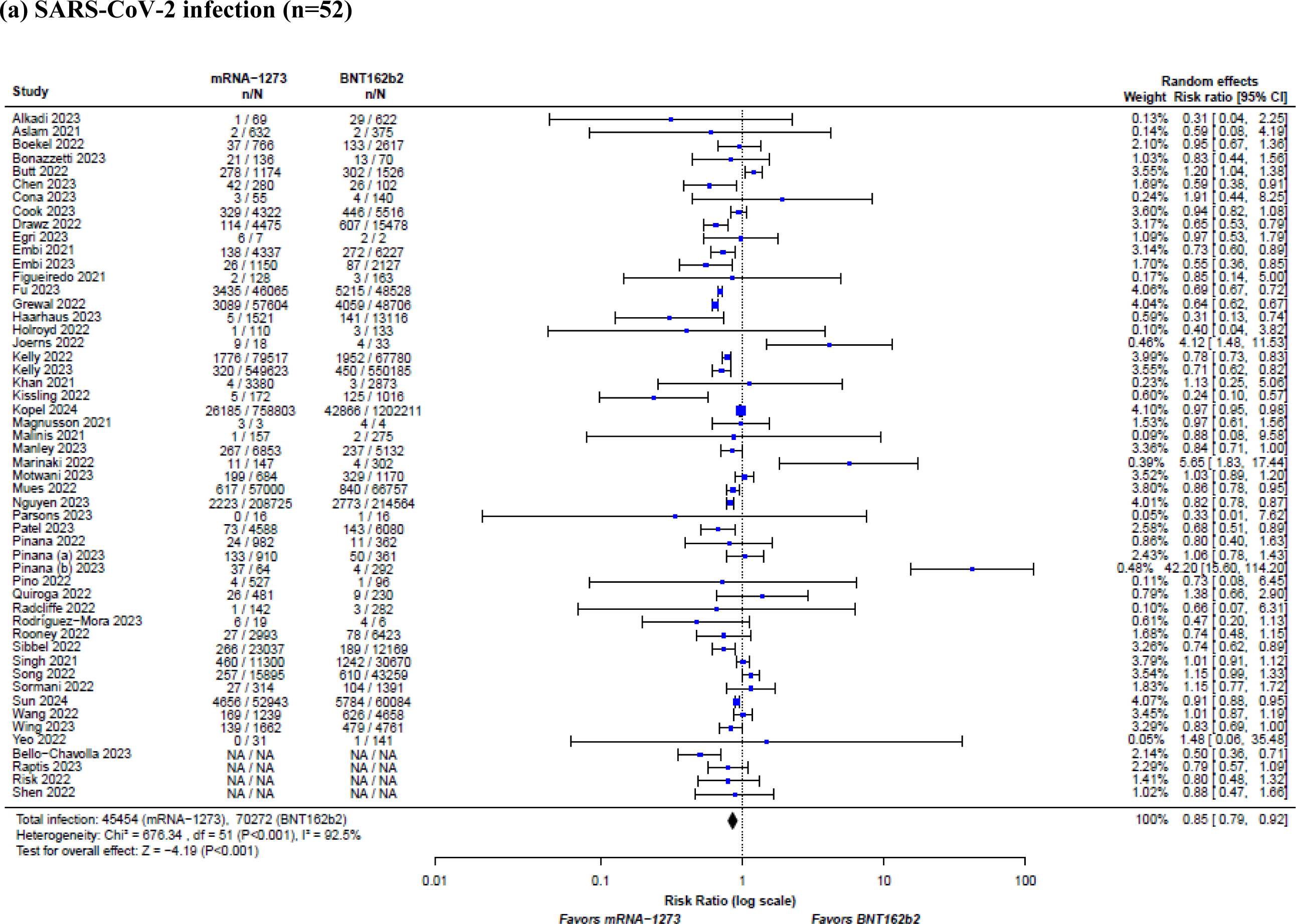
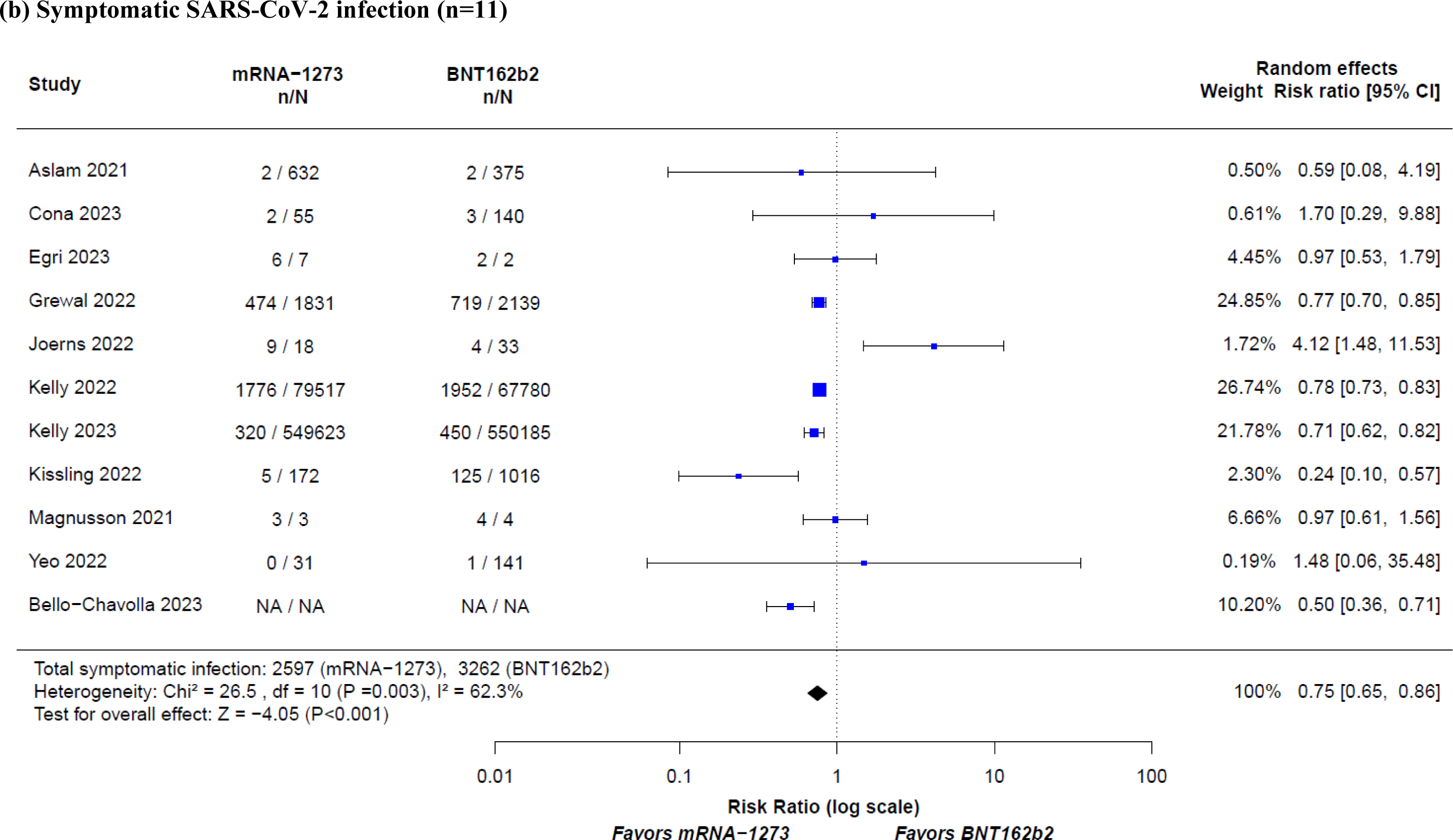
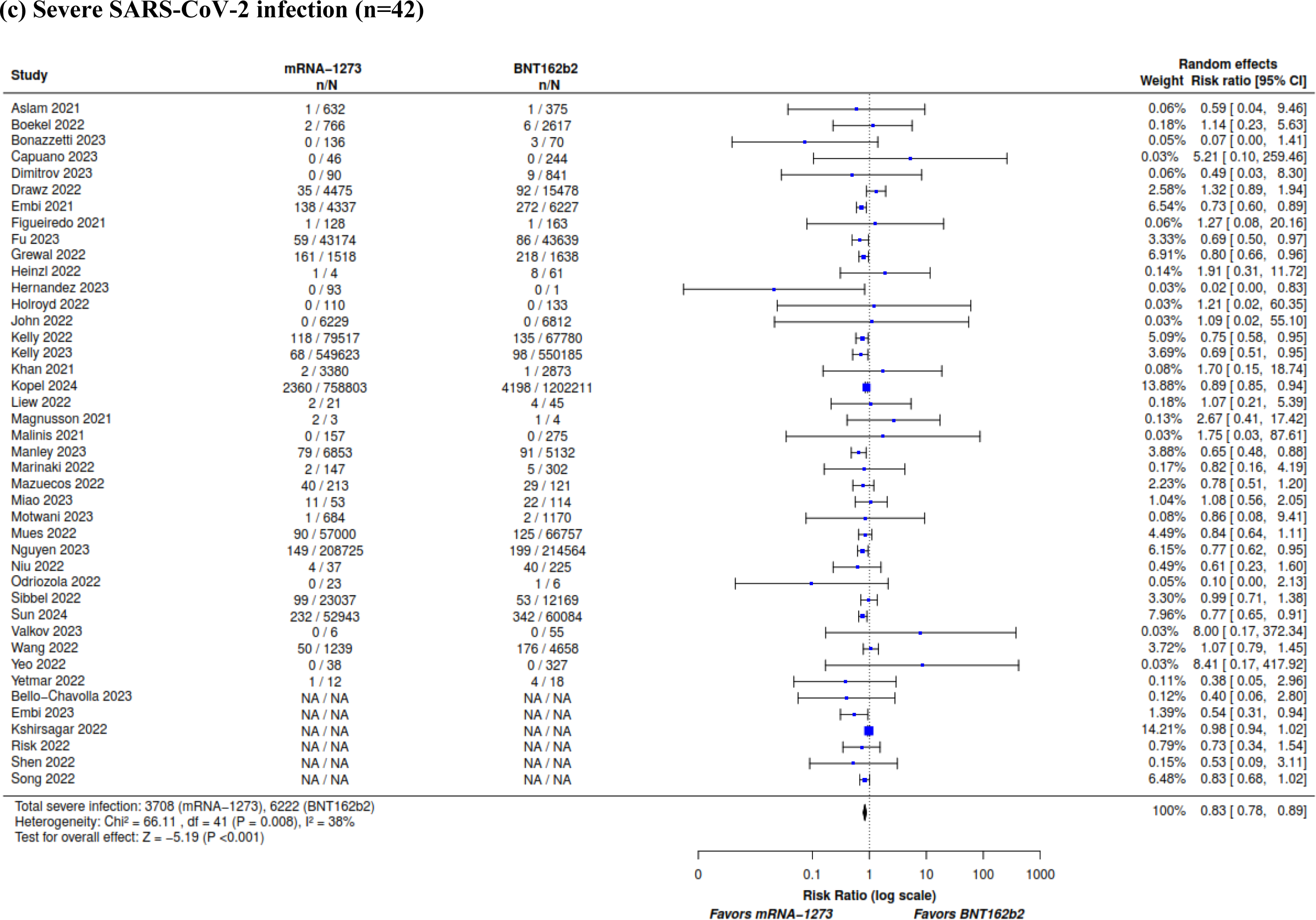
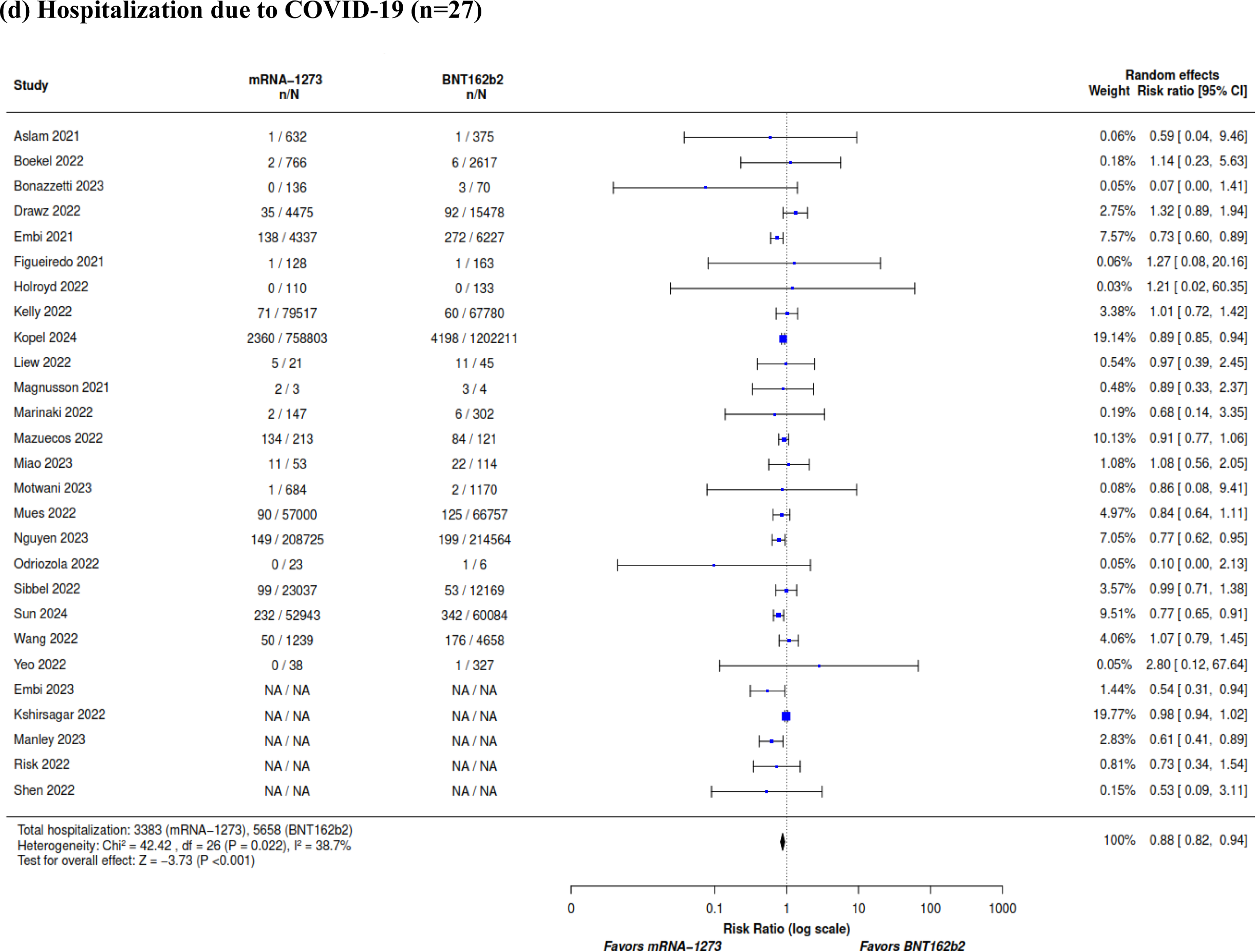
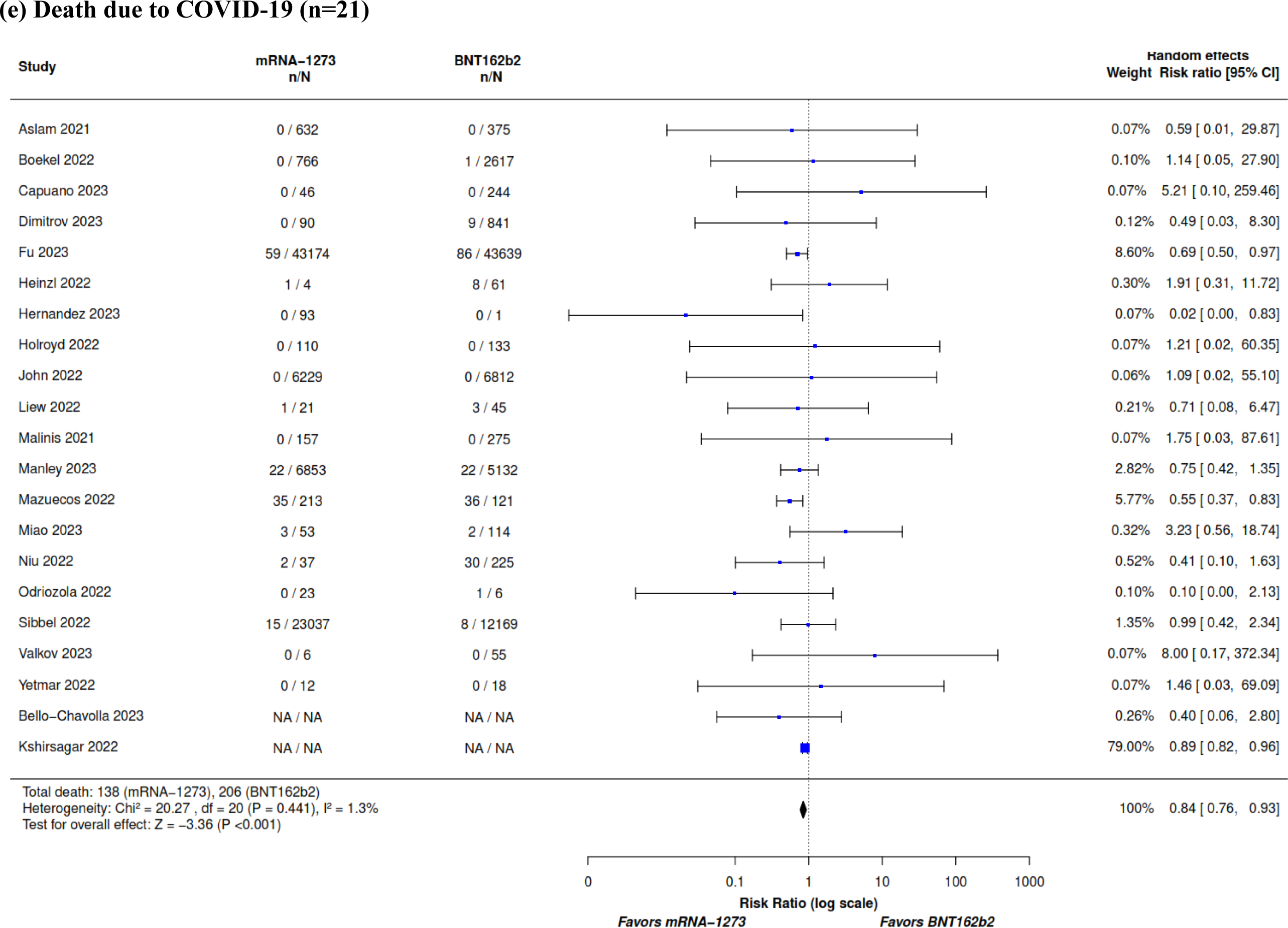
Meta-analysis results comparing the mRNA-1273 versus BNT162b2 COVID-19 vaccines in the overall population of adults with at least one medical condition^a^ by study for (a) SARS-CoV-2 infection, (b) symptomatic SARS-CoV-2 infection, (c) severe SARS-CoV-2 infection, (d) hospitalization due to COVID-19, and (e) death due to COVID-19. ^a^Autoimmune disease, solid tumor, solid organ transplant, hematologic malignancy, chronic kidney disease with and without hemodialysis, type 1 and 2 diabetes, cardiovascular disease, cerebrovascular disease, chronic liver condition, neurologic condition, chronic respiratory condition, obesity.

In 38 studies reporting the SARS-CoV-2 infection outcome in adults with CEV 1 or 2 conditions, vaccination with mRNA-1273 was associated with a significantly lower risk of SARS-CoV-2 infection compared with BNT162b2 (RR, 0.90 [95% CI, 0.84–0.97]; **Figure 4**). Between-study heterogeneity was estimated to be considerable (*I^2^*=80.2%).

**Fig. 4.**
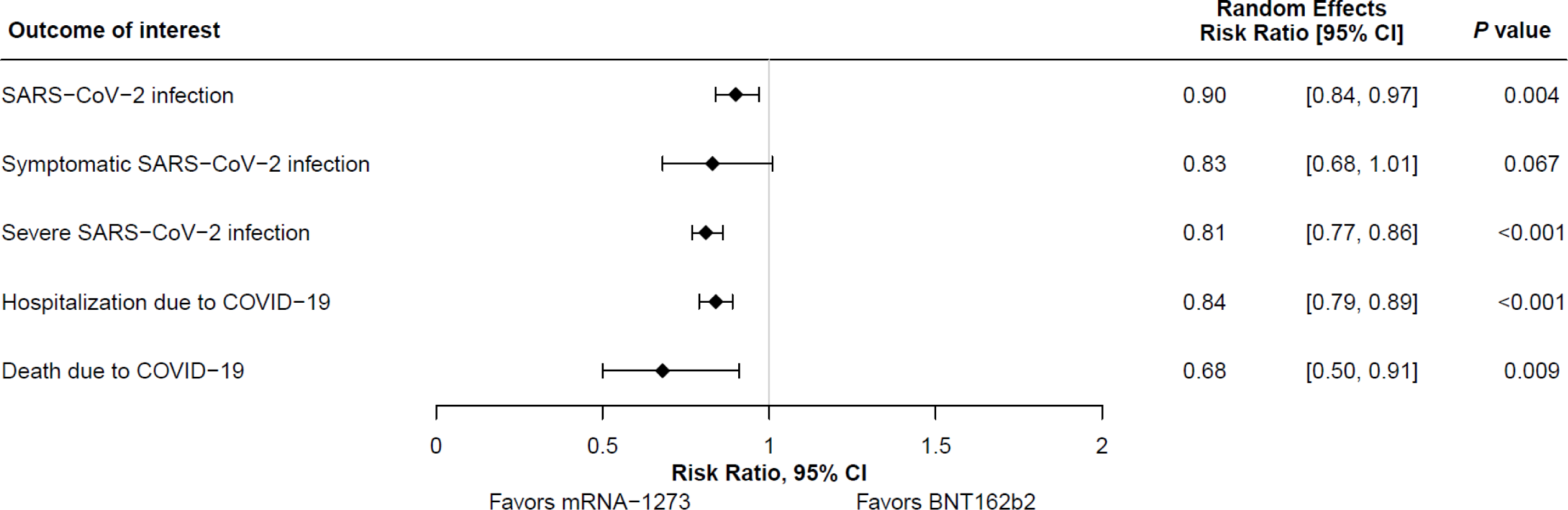
Summary of meta-analysis results on clinical effectiveness outcomes of the mRNA-1273 versus BNT162b2 COVID-19 vaccines in adults with CEV 1 and 2 conditions. CEV, clinically extremely vulnerable.

The certainty of evidence in both primary meta-analyses was very low because of inconsistency and indirectness arising from heterogeneous outcome definitions and the composition of the populations analyzed; risk of bias also contributed to very low evidence certainty in adults with CEV 1 or 2 conditions (**Table 2** and **Table 3**).

**Table 2.**
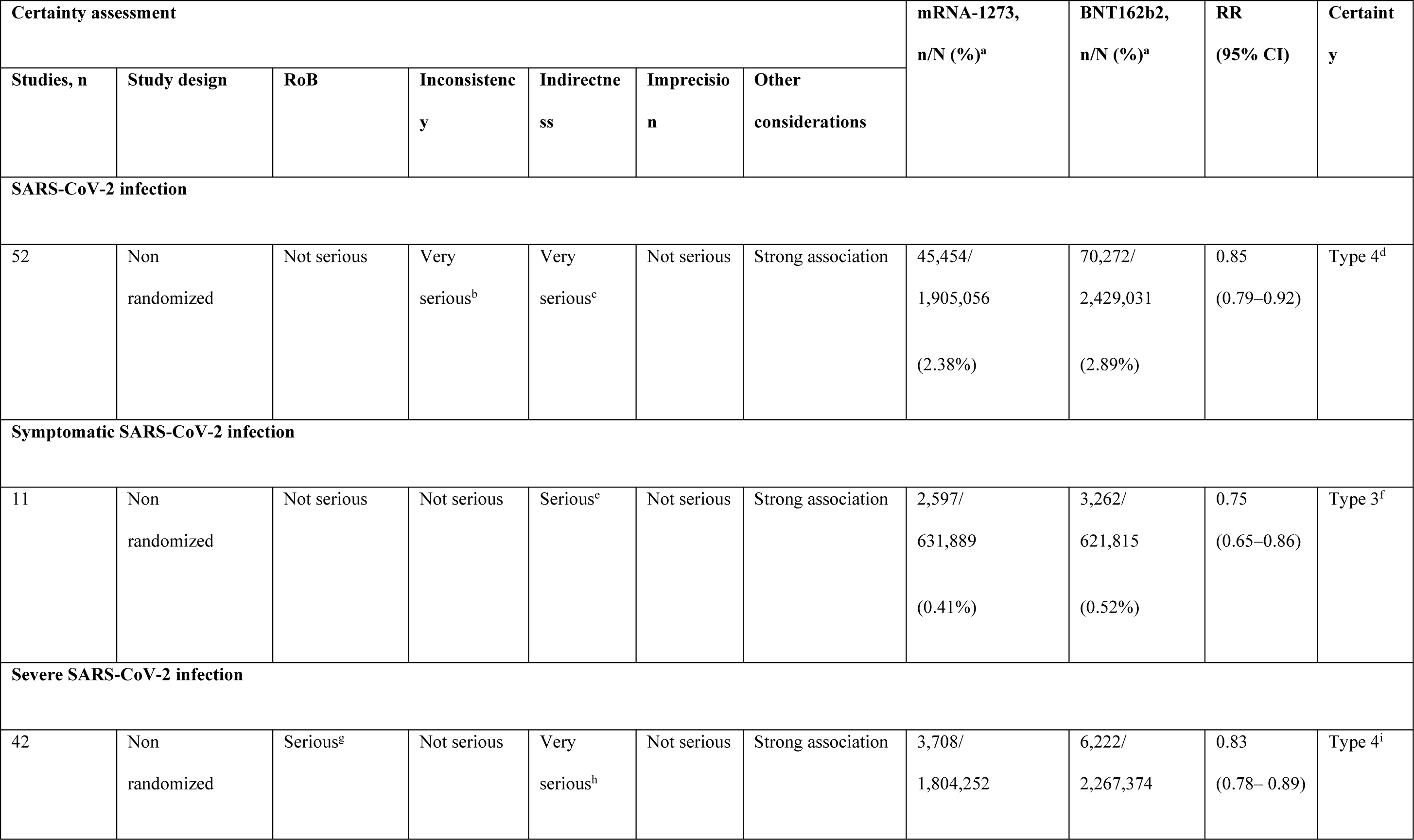
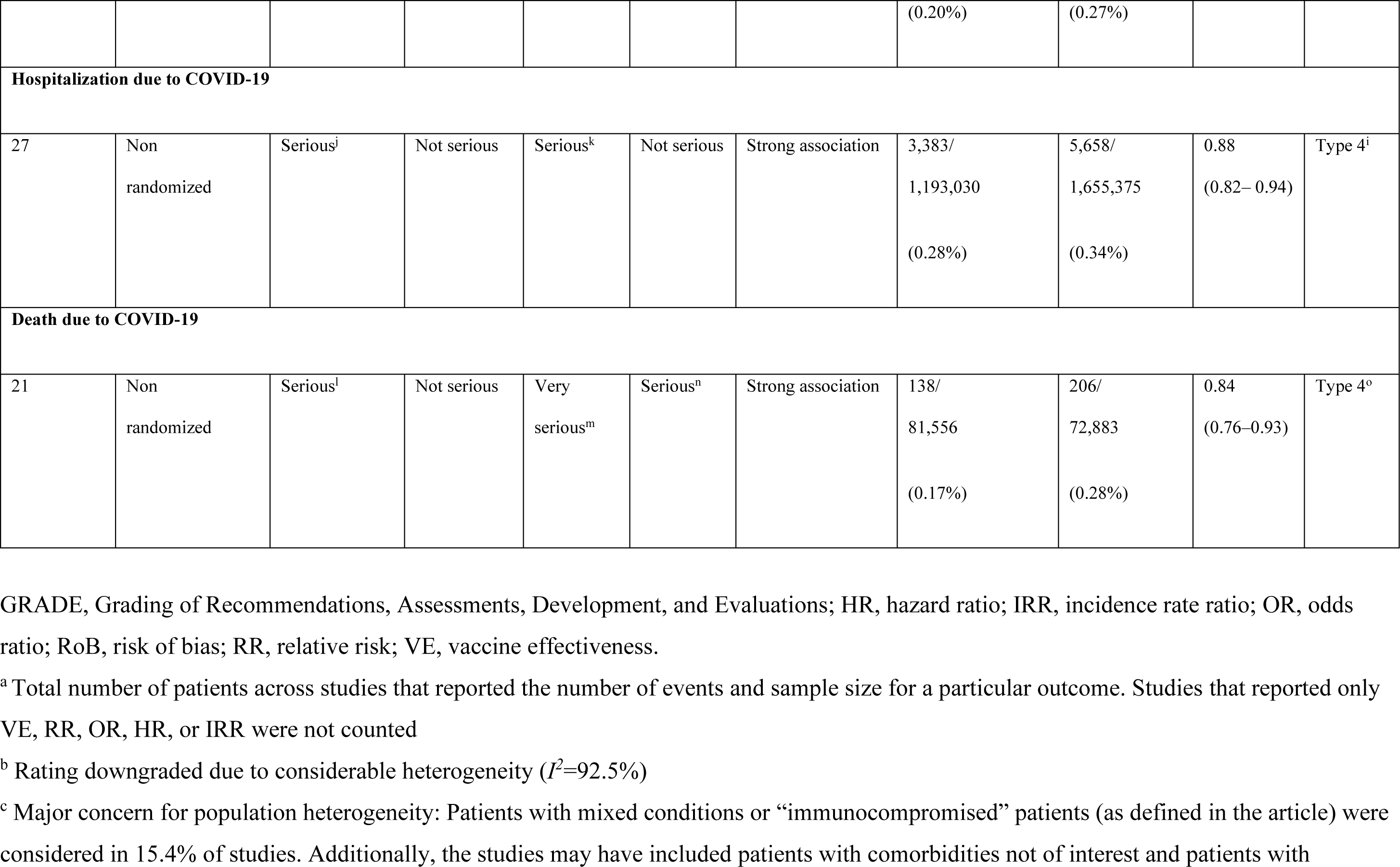
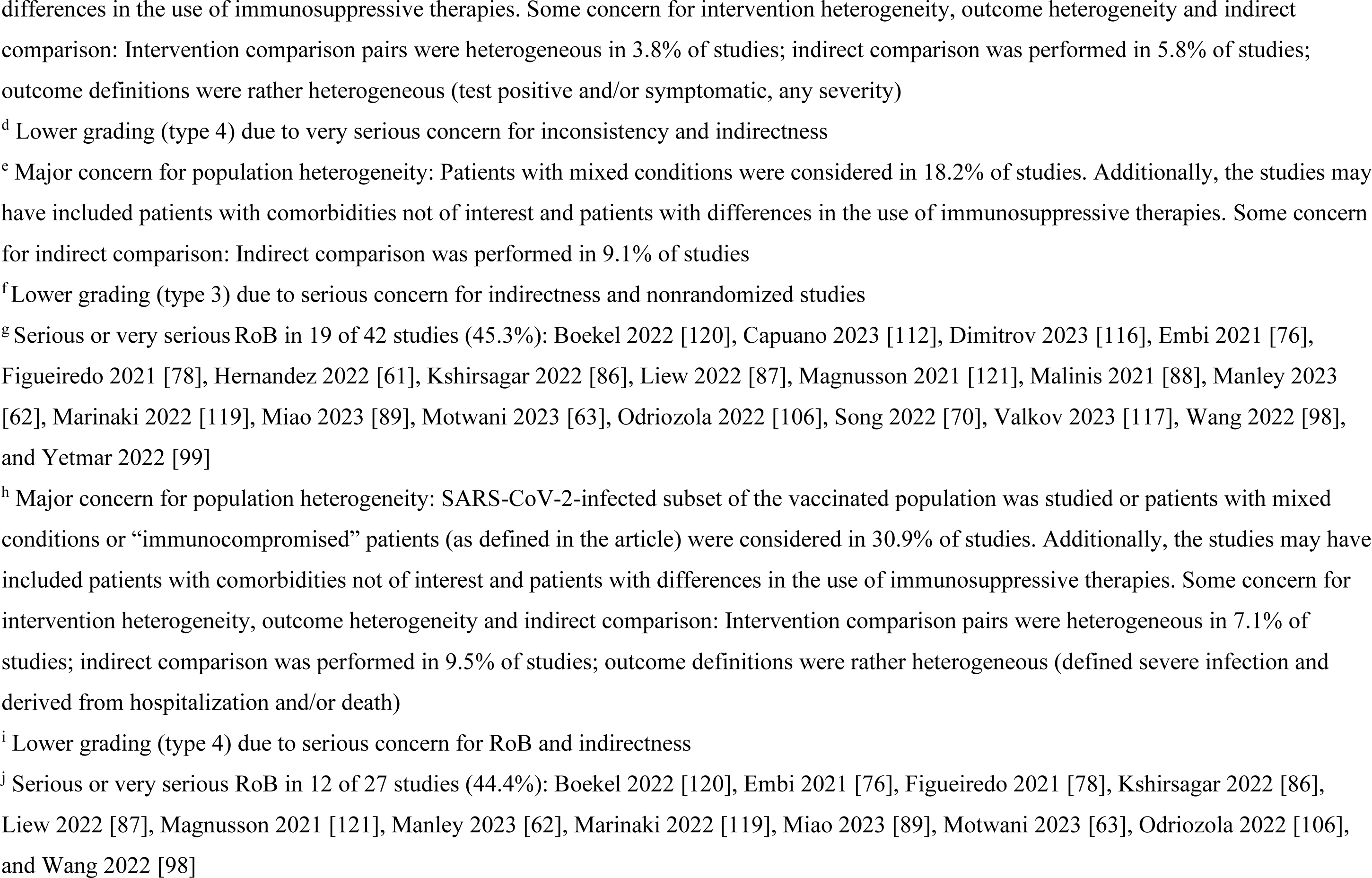
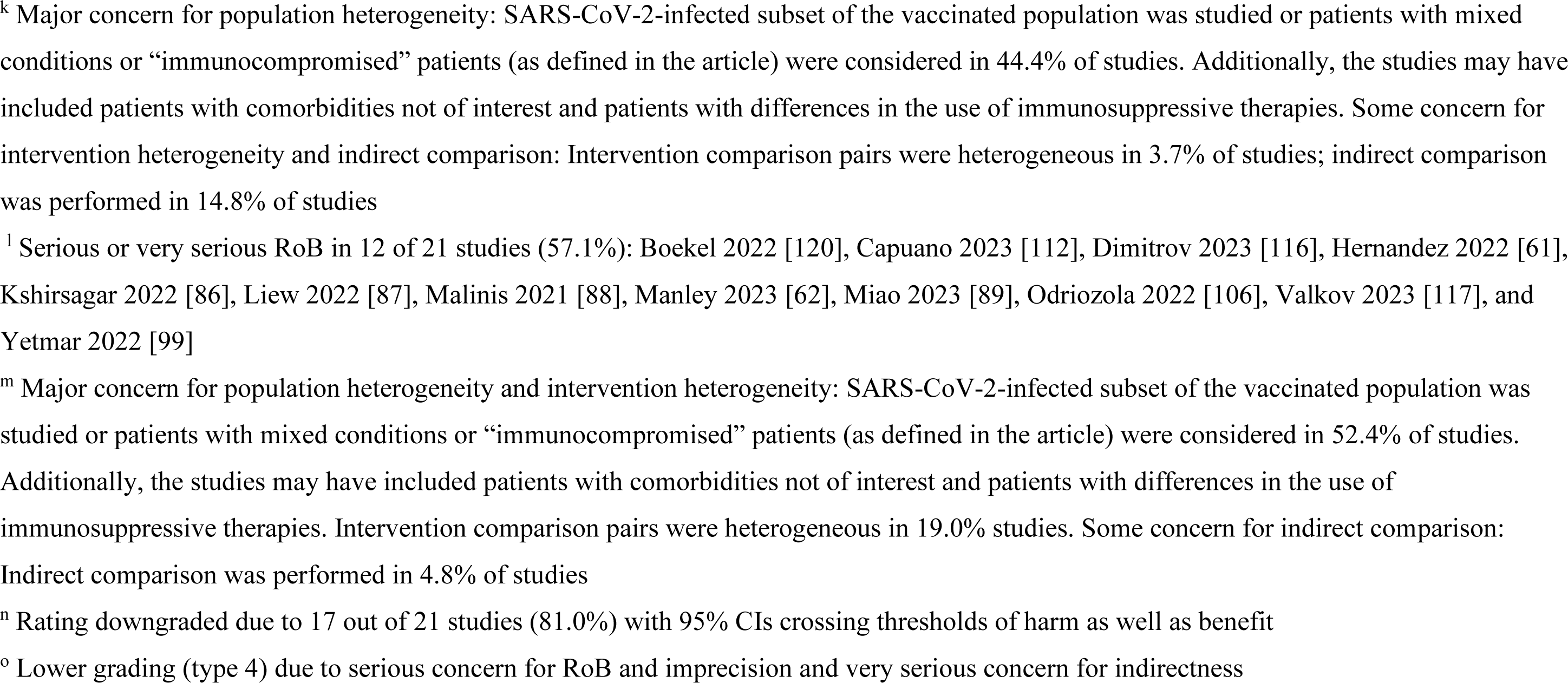
Summary of overall GRADE findings in adults with at least one underlying medical condition.

**Table 3.**
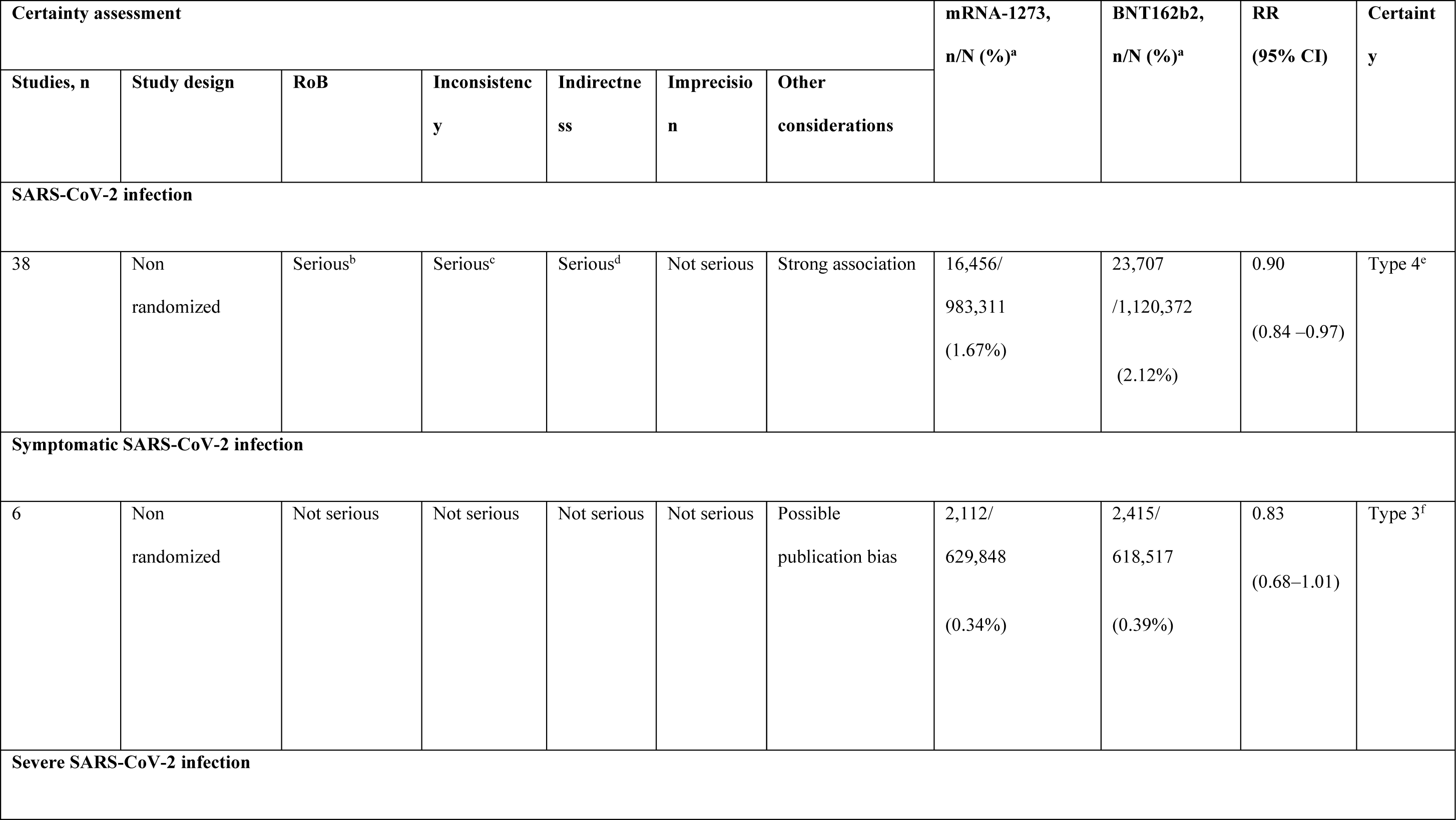
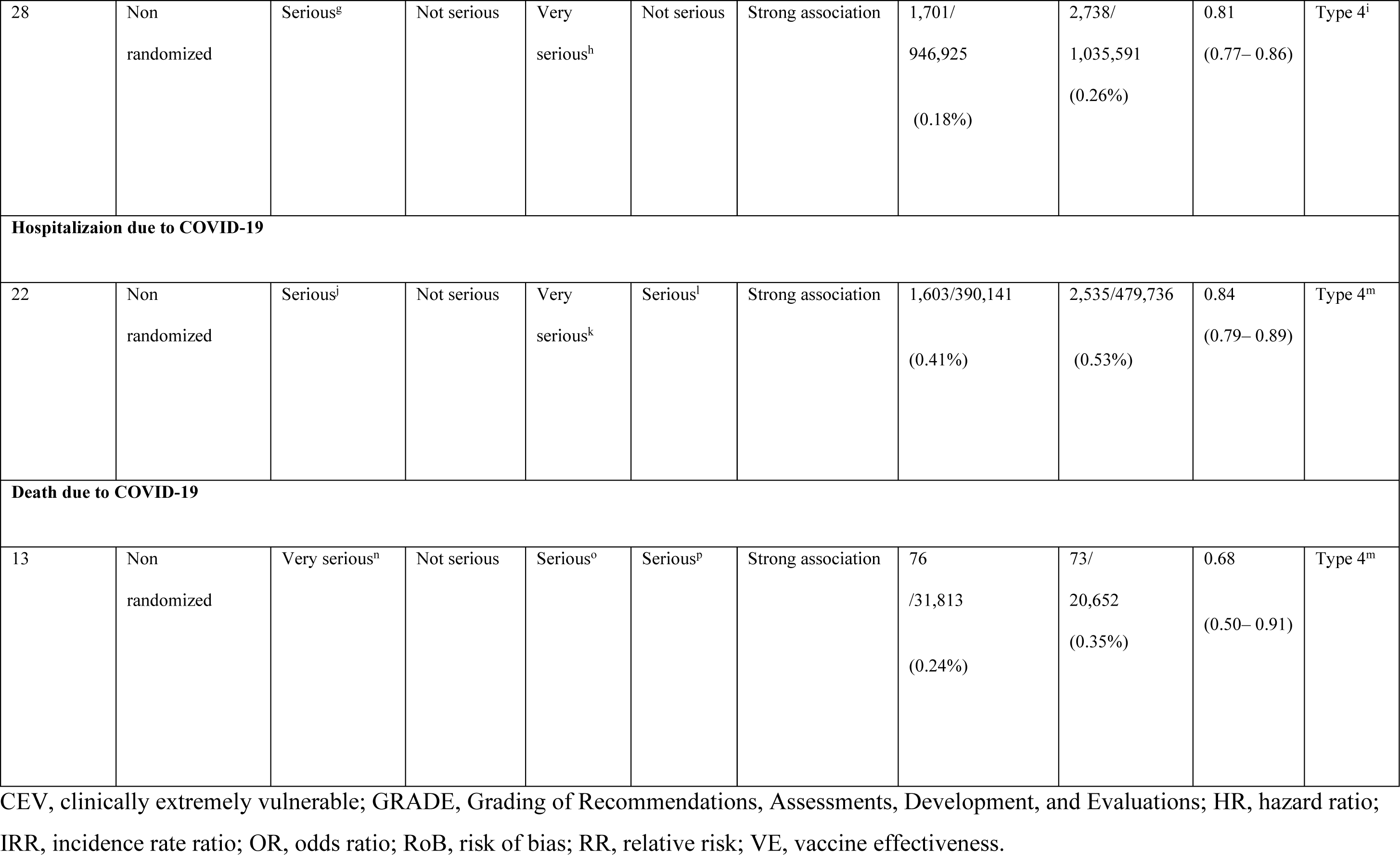
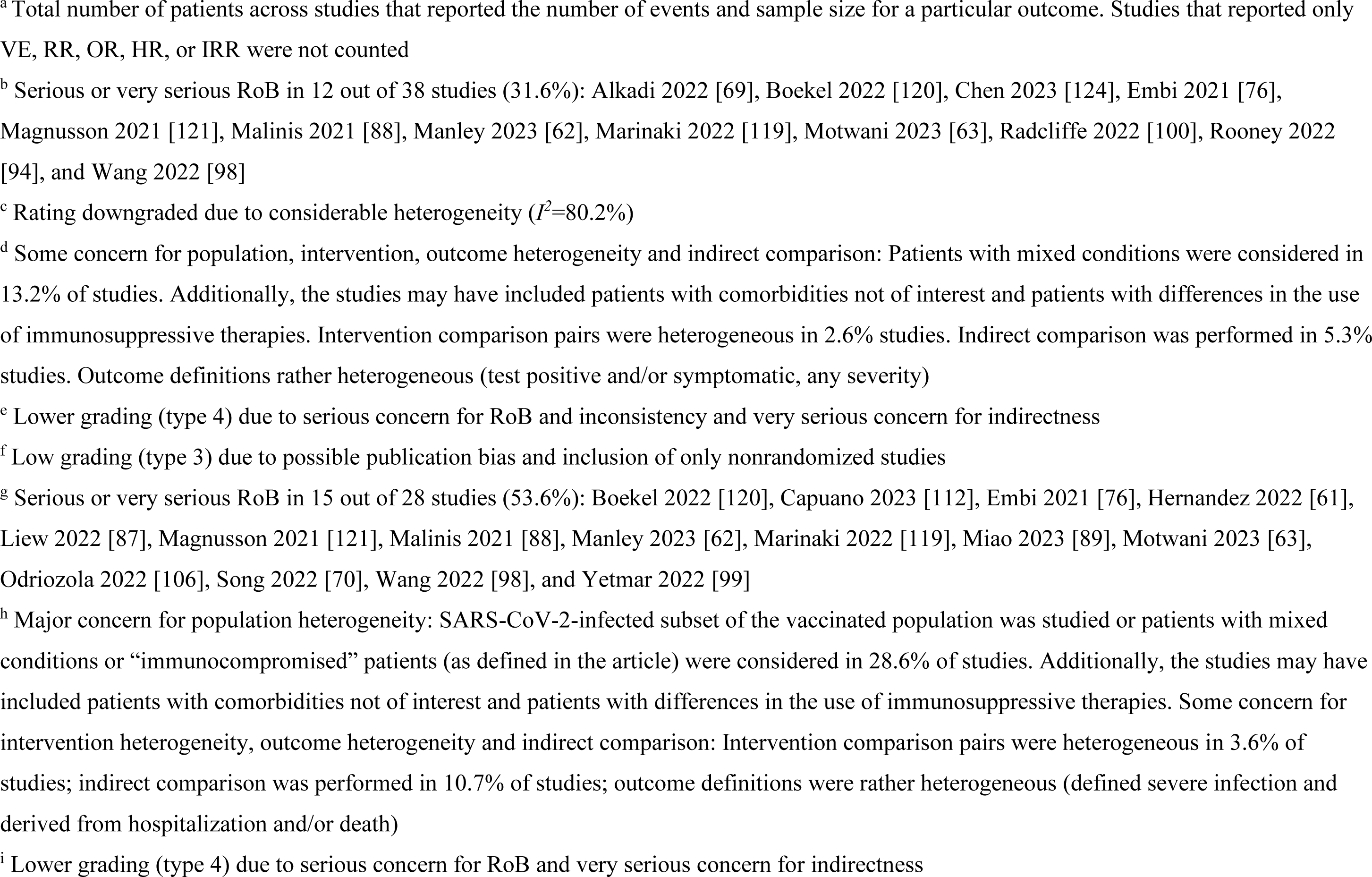
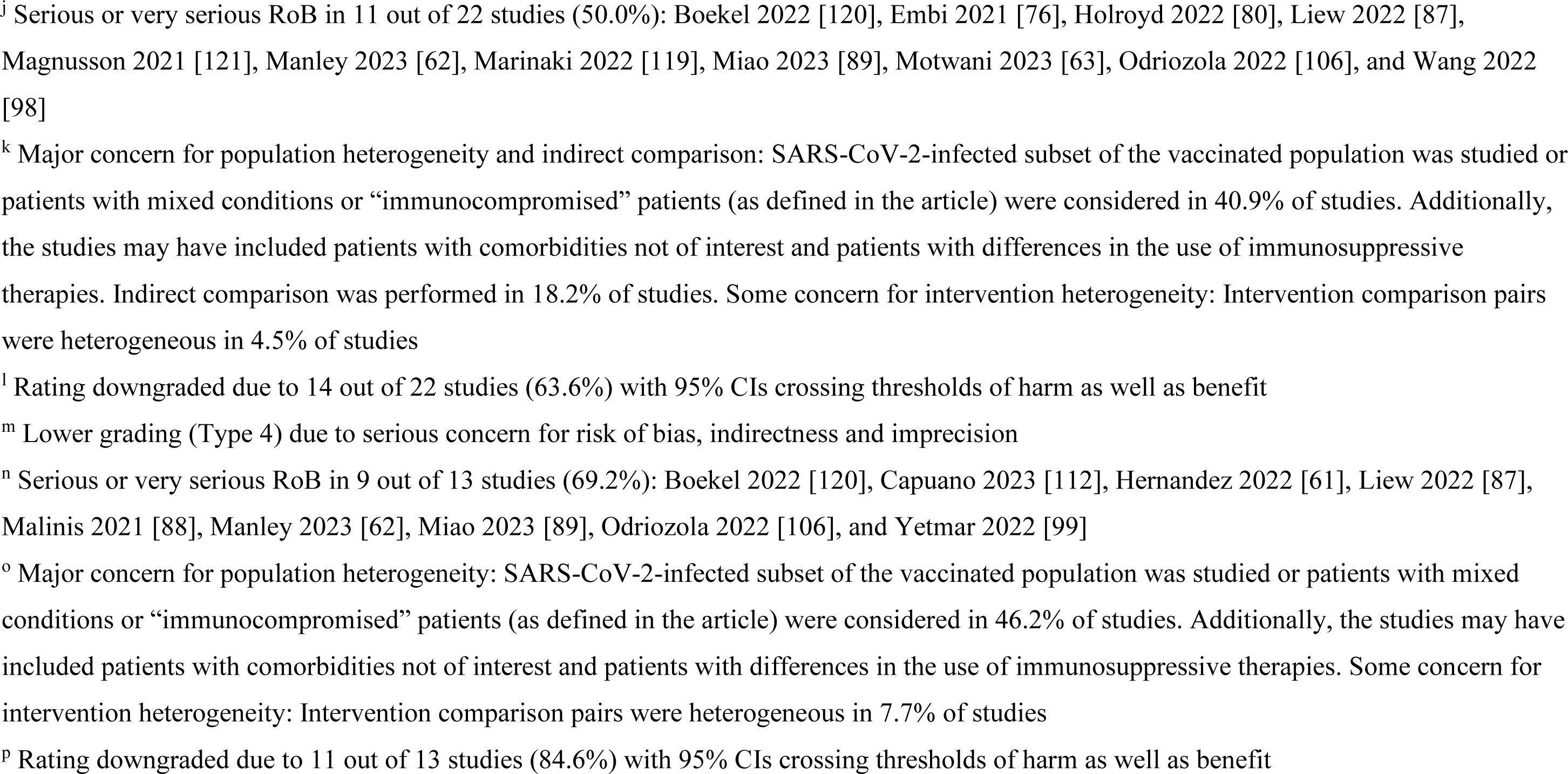
Summary of overall GRADE findings in adults with CEV 1 or 2 conditions.

| Certainty assessment |  |  |  |  |  |  | mRNA-1273,<br>n/N (%)<sup>a</sup> | BNT162b2,<br>n/N (%)<sup>a</sup> | RR<br>(95% CI) | Certainty |
| --- | --- | --- | --- | --- | --- | --- | --- | --- | --- | --- |
| Studies, n | Study design | RoB | Inconsistency | Indirectness | Imprecision | Other considerations |  |  |  |  |
| SARS-CoV-2 infection |  |  |  |  |  |  |  |  |  |  |
| 38 | Non randomized | Serious<sup>b</sup> | Serious<sup>c</sup> | Serious<sup>d</sup> | Not serious | Strong association | 16,456/<br>983,311<br>(1.67%) | 23,707<br>/1,120,372<br>(2.12%) | 0.90<br>(0.84 –0.97) | Type 4<sup>e</sup> |
| Symptomatic SARS-CoV-2 infection |  |  |  |  |  |  |  |  |  |  |
| 6 | Non randomized | Not serious | Not serious | Not serious | Not serious | Possible publication bias | 2,112/<br>629,848<br>(0.34%) | 2,415/<br>618,517<br>(0.39%) | 0.83<br>(0.68–1.01) | Type 3<sup>f</sup> |
| Severe SARS-CoV-2 infection |  |  |  |  |  |  |  |  |  |  |
| 28 | Non<br>randomized | Serious<sup>g</sup> | Not serious | Very<br>serious<sup>h</sup> | Not serious | Strong association | 1,701/<br>946,925<br>(0.18%) | 2,738/<br>1,035,591<br>(0.26%) | 0.81<br>(0.77– 0.86) | Type 4<sup>i</sup> |
| **Hospitalizaion due to COVID-19** |  |  |  |  |  |  |  |  |  |  |
| 22 | Non<br>randomized | Serious<sup>j</sup> | Not serious | Very<br>serious<sup>k</sup> | Serious<sup>l</sup> | Strong association | 1,603/390,141<br>(0.41%) | 2,535/479,736<br>(0.53%) | 0.84<br>(0.79– 0.89) | Type 4<sup>m</sup> |
| **Death due to COVID-19** |  |  |  |  |  |  |  |  |  |  |
| 13 | Non<br>randomized | Very serious<sup>n</sup> | Not serious | Serious<sup>o</sup> | Serious<sup>p</sup> | Strong association | 76<br>/31,813<br>(0.24%) | 73/<br>20,652<br>(0.35%) | 0.68<br>(0.50– 0.91) | Type 4<sup>m</sup> |

CEV, clinically extremely vulnerable; GRADE, Grading of Recommendations, Assessments, Development, and Evaluations; HR, hazard ratio; IRR, incidence rate ratio; OR, odds ratio; RoB, risk of bias; RR, relative risk; VE, vaccine effectiveness.
<sup>a</sup> Total number of patients across studies that reported the number of events and sample size for a particular outcome. Studies that reported only VE, RR, OR, HR, or IRR were not counted
<sup>b</sup> Serious or very serious RoB in 12 out of 38 studies (31.6%): Alkadi 2022 [69], Boekel 2022 [120], Chen 2023 [124], Embi 2021 [76], Magnusson 2021 [121], Malinis 2021 [88], Manley 2023 [62], Marinaki 2022 [119], Motwani 2023 [63], Radcliffe 2022 [100], Rooney 2022 [94], and Wang 2022 [98]
<sup>c</sup> Rating downgraded due to considerable heterogeneity ( $I^2=80.2\%$ )
<sup>d</sup> Some concern for population, intervention, outcome heterogeneity and indirect comparison: Patients with mixed conditions were considered in 13.2% of studies. Additionally, the studies may have included patients with comorbidities not of interest and patients with differences in the use of immunosuppressive therapies. Intervention comparison pairs were heterogeneous in 2.6% studies. Indirect comparison was performed in 5.3% studies. Outcome definitions rather heterogeneous (test positive and/or symptomatic, any severity)
<sup>e</sup> Lower grading (type 4) due to serious concern for RoB and inconsistency and very serious concern for indirectness
<sup>f</sup> Low grading (type 3) due to possible publication bias and inclusion of only nonrandomized studies
<sup>g</sup> Serious or very serious RoB in 15 out of 28 studies (53.6%): Boekel 2022 [120], Capuano 2023 [112], Embi 2021 [76], Hernandez 2022 [61], Liew 2022 [87], Magnusson 2021 [121], Malinis 2021 [88], Manley 2023 [62], Marinaki 2022 [119], Miao 2023 [89], Motwani 2023 [63], Odriozola 2022 [106], Song 2022 [70], Wang 2022 [98], and Yetmar 2022 [99]
<sup>h</sup> Major concern for population heterogeneity: SARS-CoV-2-infected subset of the vaccinated population was studied or patients with mixed conditions or “immunocompromised” patients (as defined in the article) were considered in 28.6% of studies. Additionally, the studies may have included patients with comorbidities not of interest and patients with differences in the use of immunosuppressive therapies. Some concern for intervention heterogeneity, outcome heterogeneity and indirect comparison: Intervention comparison pairs were heterogeneous in 3.6% of studies; indirect comparison was performed in 10.7% of studies; outcome definitions were rather heterogeneous (defined severe infection and derived from hospitalization and/or death)
<sup>i</sup> Lower grading (type 4) due to serious concern for RoB and very serious concern for indirectness
<sup>j</sup> Serious or very serious RoB in 11 out of 22 studies (50.0%): Boekel 2022 [120], Embi 2021 [76], Holroyd 2022 [80], Liew 2022 [87], Magnusson 2021 [121], Manley 2023 [62], Marinaki 2022 [119], Miao 2023 [89], Motwani 2023 [63], Odriozola 2022 [106], and Wang 2022 [98]
<sup>k</sup> Major concern for population heterogeneity and indirect comparison: SARS-CoV-2-infected subset of the vaccinated population was studied or patients with mixed conditions or “immunocompromised” patients (as defined in the article) were considered in 40.9% of studies. Additionally, the studies may have included patients with comorbidities not of interest and patients with differences in the use of immunosuppressive therapies. Indirect comparison was performed in 18.2% of studies. Some concern for intervention heterogeneity: Intervention comparison pairs were heterogeneous in 4.5% of studies
<sup>l</sup> Rating downgraded due to 14 out of 22 studies (63.6%) with 95% CIs crossing thresholds of harm as well as benefit
<sup>m</sup> Lower grading (Type 4) due to serious concern for risk of bias, indirectness and imprecision

Subgroup meta-analyses of studies reporting SARS-CoV-2 infection by vaccine regimen, age group, and SARS-CoV-2 variant were generally similar to the primary results in adults with at least one underlying medical condition (**Figure 5a**) and in adults with CEV 1 or 2 conditions (**Figure 5b**). Trends towards reduced risk of SARS-CoV-2 infection among patients who received mRNA-1273 versus BNT162b2 were observed across all of the subgroups analyzed. In particular, mRNA-1273 was associated with a statistically significant reduction in infection risk in both the SARS-CoV-2 Delta and Omicron variant subgroups in adults with at least one underlying medical condition and CEV 1 and 2 conditions.

**Fig. 5.**
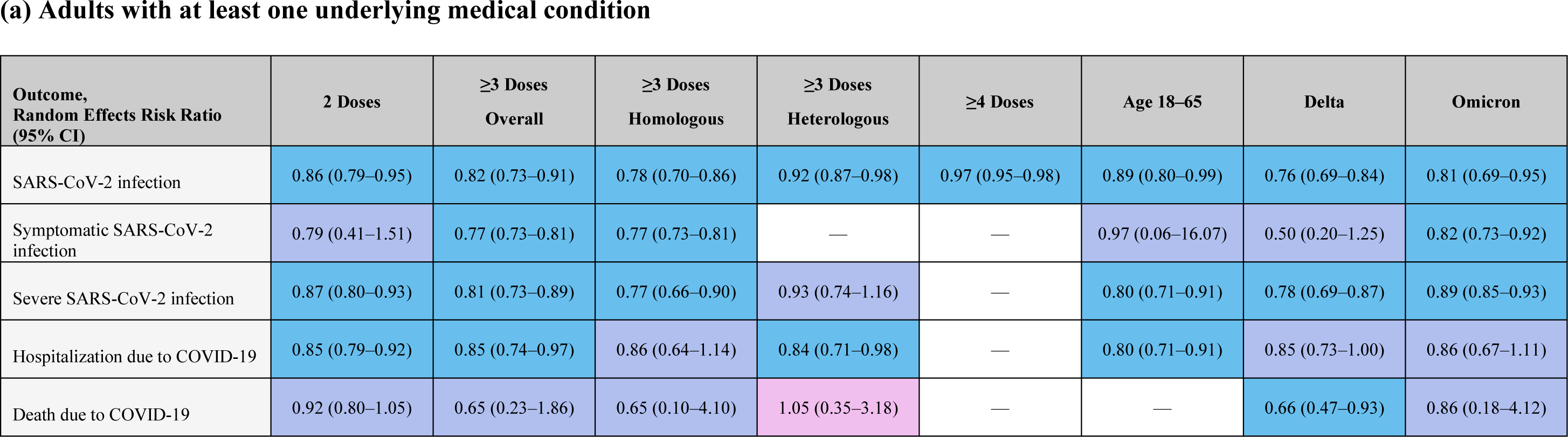
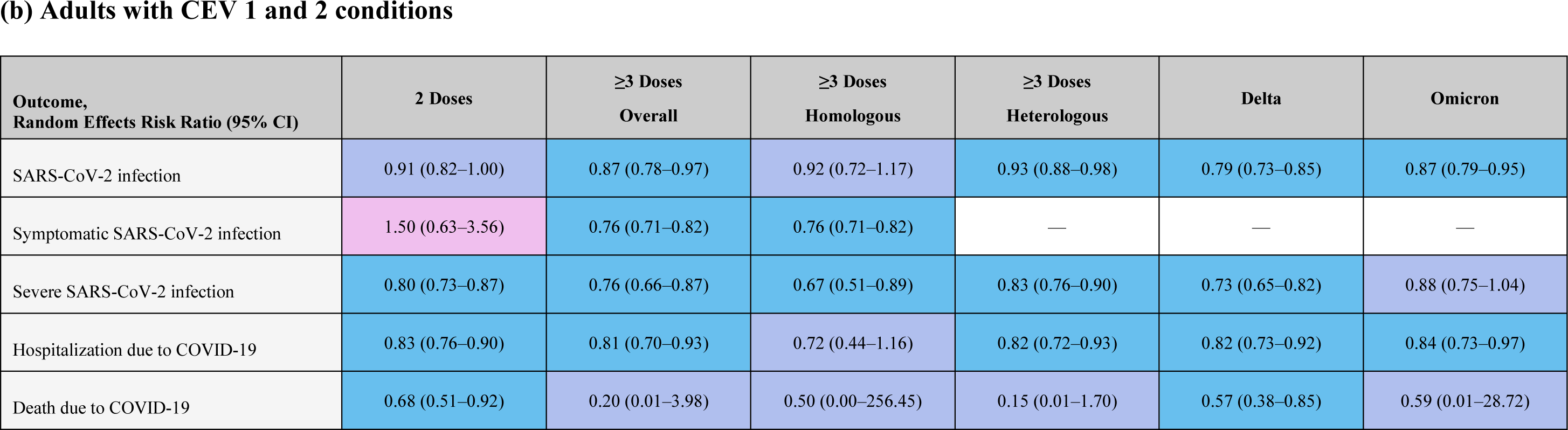
Summary of subgroup meta-analysis results on clinical effectiveness outcomes of the mRNA-1273 versus BNT162b2 COVID-19 vaccines in adults with (a) at least one underlying medical condition^a^ and (b) CEV 1 and 2 conditions. ^a^Autoimmune disease, solid tumor, solid organ transplant, hematologic malignancy, chronic kidney disease with and without hemodialysis, type 1 and 2 diabetes, cardiovascular disease, cerebrovascular disease, chronic liver condition, neurologic condition, chronic respiratory condition, obesity. Blue shading represents statistically significant RR in favor of mRNA-1273 over BNT162b2, purple shading represents statistically nonsignificant RR, and pink shading represents statistically nonsignificant RR in favor of BNT162b2 over mRNA-1273. CEV, clinically extremely vulnerable; RR, risk ratio.

### Symptomatic SARS-CoV-2 infection

A total of 11 studies reporting symptomatic SARS-CoV-2 infection were included in the meta-analysis of adults with at least one underlying medical condition. In this population, vaccination with mRNA-1273 was associated with a significantly lower risk of infection compared with BNT162b2 (RR, 0.75 [95% CI, 0.65–0.86]; **Figure 2**; **Figure 3b**). Heterogeneity between studies was estimated to be substantial (*I^2^*=62.3%).

Meta-analysis of 6 studies reporting data for adults with CEV 1 or 2 conditions found that vaccination with mRNA-1273 was associated with a trend towards lower risk of symptomatic infection compared with vaccination with BNT162b2 (RR, 0.83 [95% CI, 0.68–1.01]; **Figure 4**); however, this result was not statistically significant. There was substantial heterogeneity between studies (*I^2^*=62.2%).

The certainty of evidence in both primary meta-analyses was low because only nonrandomized studies were included, and because of indirectness resulting from heterogeneity in the composition of the population of adults with at least one underlying medical condition (**Table 2** and **Table 3**).

When analyzed by vaccine regimen, age group, and SARS-CoV-2 variant subgroups, trends toward reduced risk of symptomatic SARS-CoV-2 infection with mRNA-1273 vaccination versus BNT162b vaccination were observed in adults with at least one underlying medical condition (**Figure 5a**) and in adults with CEV 1 or 2 conditions (**Figure 5b**). All subgroups analyzed were consistent with the primary analyses, except for adults with CEV 1 or 2 conditions who received 2 doses (RR, 1.50 [95% CI, 0.63–3.56]; *I^2^*=56.7%).

### Severe SARS-CoV-2 infection

Meta-analysis of 42 studies reporting the severe SARS-CoV-2 infection outcome found that vaccination with mRNA-1273 was associated with a significantly lower risk of severe SARS-CoV-2 infection compared with BNT162b2 in adults with at least one underlying medical condition (RR, 0.83 [95% CI, 0.78–0.89]; **Figure 2** and **Figure 3c**). Heterogeneity was estimated to likely be not important between studies (*I^2^*=38.0%).

Meta-analysis of 28 studies reporting the severe SARS-CoV-2 infection outcome in adults with CEV 1 or 2 conditions found that vaccination with mRNA-1273 was associated with a significantly lower risk of severe SARS-CoV-2 infection compared with vaccination with BNT162b2 (RR, 0.81 [95% CI, 0.77–0.86]; **Figure 4**). No heterogeneity between the studies was observed (*I^2^*=0%).

The certainty of evidence in both primary meta-analyses was very low because of risk of bias and indirectness resulting primarily from heterogeneity in the composition of the populations analyzed (**Table 2** and **Table 3**).

Subgroup meta-analyses of studies reporting severe SARS-CoV-2 infection by vaccine regimen, age group, and SARS-CoV-2 variant were generally similar to the primary results in adults with at least one underlying medical condition (**Figure 5a**) and in patients with CEV 1 or 2 conditions (**Figure 5b**). Trends towards reduced risk of severe SARS-CoV-2 infection among patients who received mRNA-1273 versus BNT162b2 were observed in all of the subgroups analyzed, including the Delta and Omicron variant subgroups.

### Hospitalization due to COVID-19

Meta-analysis of 27 studies reporting hospitalization due to COVID-19 found that vaccination with mRNA-1273 was associated with a significantly lower risk of hospitalization compared with BNT162b2 in adults with at least one underlying medical condition (RR, 0.88 [95% CI, 0.82–0.94]; **Figure 2** and **Figure 3d**). Heterogeneity between studies was estimated to likely be not important (*I^2^*=38.7%).

In 22 studies reporting hospitalization due to COVID-19 in adults with CEV 1 or 2 conditions, vaccination with mRNA-1273 was associated with a significantly lower risk of hospitalization compared with BNT162b2 (RR, 0.84 [95% CI, 0.79–0.89]; **Figure 4**). No heterogeneity between studies was observed (*I^2^*=0%).

The certainty of evidence in both primary meta-analyses was very low because of risk of bias and indirectness resulting primarily from heterogeneity in the composition of the population analyzed (**Table 2** and **Table 3**).

Results from subgroup meta-analyses of studies reporting hospitalization due to COVID-19 by vaccine regimen, age group, and SARS-CoV-2 variant were generally similar to the primary results in adults with at least one underlying medical condition (**Figure 5a**) and in adults with CEV 1 or 2 conditions (**Figure 5b**). Trends towards reduced risk of hospitalization among patients who received mRNA-1273 versus BNT162b2 were observed across all of the subgroups analyzed, including the Delta and Omicron SARS-CoV-2 variant subgroups.

### Death due to COVID-19

Meta-analysis of 21 studies reporting the death due to COVID-19 outcome found that vaccination with mRNA-1273 was associated with a significantly lower risk of death due to COVID-19 compared with vaccination with BNT162b2 in adults with at least one underlying medical condition (RR, 0.84 [95% CI, 0.76–0.93]; **Figure 2** and **Figure 3e**). Heterogeneity between studies was estimated to likely be not important (*I^2^*=1.3%).

In 13 studies reporting death due to COVID-19 in adults with CEV 1 or 2 conditions, vaccination with mRNA-1273 was associated with a significantly lower risk of death due to COVID-19 compared with BNT162b2 (RR, 0.68 [95% CI, 0.50–0.91]; **Figure 4**). No heterogeneity between studies was observed (*I^2^*=0%).

The certainty of evidence in both primary meta-analyses was very low because of risk of bias, imprecision, and indirectness resulting primarily from the heterogeneity in the composition of the populations analyzed (**Table 2** and **Table 3**).

When analyzed by vaccine regimen, age group, and SARS-CoV-2 variant subgroups, trends toward reduced risk of death due to COVID-19 with mRNA-1273 vaccination versus BNT162b vaccination were observed in adults with at least one underlying medical condition (**Figure 5a**) and in adults with CEV 1 or 2 conditions (**Figure 5b**). All subgroups analyzed, including the Delta and Omicron SARS-CoV-2 variant subgroups were consistent with the primary analyses, except in adults with at least one underlying medical condition who received ≥3 heterologous vaccine doses (RR, 1.05 [95% CI, 0.35–3.18]; *I^2^*=23.8%).

## DISCUSSION

In this meta-analysis of 65 studies comprising more than 9 million vaccinated adults with at least one underlying medical condition putting them at risk for developing severe COVID-19, we found that vaccination with mRNA-1273 was associated with a significantly lower risk of asymptomatic and symptomatic SARS-CoV-2 infections, severe SARS-CoV-2 infection, hospitalization due to COVID-19, and death due to COVID-19 compared with BNT162b2. Our meta-analysis also consistently demonstrated across dosing regimens and in immunocompromised patients and population subgroups with selected high-risk comorbidities that vaccination with mRNA-1273 led to improved outcomes. The observed trend, that the benefit associated with mRNA-1273 vaccination compared with BNT162b2 vaccination in reducing the risk of the severe COVID-19 outcomes (severe SARS-CoV-2 infection, hospitalization due to COVID-19, and death due to COVID-19) was higher among adults with CEV groups 1 or 2 conditions than the larger overall population of adults with at least one underlying medical condition, suggests that the importance of choice of mRNA vaccine increases with the severity of the underlying conditions. For the immunocompromised population (ie, adults with CEV 1 and 2 conditions), the findings of our meta-analysis confirm results of a previous GRADE meta-analysis in the same population which primarily considered studies on the primary vaccine series [12], as well as the findings of a GRADE meta-analysis in the vulnerable population of older adults, many of whom are likely to have similar comorbidities [4,13]. As it would be impracticable to generate comparative RCT evidence for mRNA-1273 versus BNT162b2 in the heterogeneous population of patients with medical conditions and in particular immunocompromised patients, SLR and GRADE meta-analysis of observational studies represents the most robust source of information [126] for decision makers in the vaccine ecosystem helping to protect patients with underlying medical conditions against severe COVID-19.

Patients at high risk for developing severe COVID-19 illness based on their medical history may have more than one underlying medical condition influencing risk, as has been recognized in previous estimates of the global at-risk population [4]. Because no assumptions were made about a single primary medical condition in the populations of our primary analyses, our finding that vaccination with mRNA-1273 was associated with reduced risk of severe COVID-19 in adults with at least one underlying medical condition and patients who are immunocompromised is relevant also to patients who may have multiple and varied comorbidities. Interpreting results for patient subgroups defined by groups of medical conditions and individual diseases is challenging because smaller sample sizes and disease-specific factors (eg, use of immunosuppressive medications in some patients with autoimmune disease) may mask differences in VE between COVID-19 mRNA vaccines.

We observed a considerable amount of heterogeneity between studies for the SARS-CoV-2 infection and symptomatic SARS-CoV-2 infection outcomes but not important or no heterogeneity for the severe SARS-CoV-2 infection, hospitalization due to COVID-19, and death due to COVID-19 outcomes. The observed heterogeneity in this meta-analysis also occurred to a lesser extent than in the previous meta-analyses of Kavikondala, et al [13] and Wang, et al [12]. Among the factors that may be driving the observed heterogeneity between studies are differences in study populations, statistical approaches employed, outcome definitions (eg, for severe COVID-19), the post-vaccination timepoints analyzed, and vaccination schedules. Subgroup analyses to account for differences in age, dosing regimens, and SARS-CoV-2 variants did not reduce the observed heterogeneity. However, the extensive subgroup analyses conducted were consistent with the primary results and thus confirm the robustness of our main findings.

At the current stage of the COVID-19 pandemic, in which there are high levels of immunity in the general population and with Omicron being the predominant circulating SARS-CoV-2 variant [1,2], the public health focus has shifted to preventing severe COVID-19 in vulnerable populations through vaccination [5]. In our Omicron subgroup analysis, there was a statistically significant reduction in risk favoring mRNA-1273 for overall, symptomatic, and severe SARS-CoV-2 infection outcomes in adults with at least one underlying medical condition and overall SARS-CoV-2 infection and hospitalization due to COVID-19 in adults with CEV 1 or 2 conditions. All remaining outcomes, except for symptomatic SARS-CoV-2 infection in adults with CEV 1 or 2 conditions, for which no studies were reported, numerically favored mRNA-1273 over BNT162b2 for the Omicron subgroup. The latter findings could be attributed to relatively small numbers of studies included in the meta-analysis of the Omicron subgroup, especially for severe events, and the heterogeneity of the target population. As data are still accumulating in the Omicron setting, statistically significant differences in VE may become apparent. However, the Omicron subgroup analysis confirms the overall findings of the main analysis, and thus suggests applicability of the findings of this meta-analysis for current and future vaccine decision-making.

A key limitation of our study is that no RCTs or head-to-head trials were identified for inclusion in our evidence synthesis. Our meta-analysis was based on observational studies, which are considered in the hierarchy of evidence to be of lower quality, resulting in a low level of certainty per GRADE. While all of the included studies were observational and considered to have some risk of bias, approximately half of the studies were considered to have a serious risk of bias or reported insufficient information to evaluate risk of bias, limiting the feasibility of sensitivity analyses. However, we conducted several subgroup analyses and found that estimates of VE were consistent with the primary analyses across all COVID-19 outcomes studied. Availability of higher quality observational studies would further add to the evidence on comparative COVID-19 VE. Many of the studies identified for this meta-analysis did not specify the SARS-CoV-2 variant reported, or authors defined variants differently. For example, Hernandez et al, reported data for Alpha, Delta, and Omicron SARS-CoV-2 infections [61] and Pinana et al, reported data for Delta SARS-CoV-2 infections only [107], despite the studies having been conducted in the same countries and during similar time periods. Most studies reported data from all vaccinated patients; however, some studies considered all infected and vaccinated patients, further contributing to the heterogeneity of the meta-analysis population. Finally, patients may have had multiple medical conditions, which makes interpreting the results presented in the **Supplementary Material** in patients grouped by medical conditions challenging.

The primary strength of our evidence synthesis is that it is, to our knowledge, the first meta-analysis to provide comparative effectiveness results for the two available COVID-19 mRNA vaccines in adults with underlying medical conditions at high risk for developing severe COVID-19 disease across original/ancestral-containing primary series and booster vaccination up until and including Omicron-containing bivalent original/B4-5 mRNA vaccines. We used broad search terms and robust SLR methodology. By searching the MEDLINE, Embase and Cochrane databases and crosschecking results with other previously published SLRs and meta-analyses, we developed a global analysis population of patients from different stages of the pandemic with multiple high-risk medical conditions represented. Original study authors were contacted to provide clarification on published data where necessary, enhancing the quality of data included in our analysis. Of all the base-case meta-analyses tested, publication bias was only suspected in adults with CEV 1 and 2 conditions for the symptomatic SARS-CoV-2 outcome, further increasing the strength of our study. In addition, using advanced meta-analysis methodology applied in other meta-analyses of COVID-19 vaccination [28,34] allowed inclusion of both studies reporting event and participant numbers by vaccine arm as well as studies reporting only VE. Notably, due to the lack of head-to-head data, our use of observational studies reporting VE data for each vaccine to compare VE between the two COVID-19 mRNA vaccines is supported by a similar analysis by the CDC Influenza Division [127].

In conclusion, vaccination with mRNA-1273 was associated with significantly lower risks of SARS-CoV-2 infection and severe COVID-19 illness in patients with underlying medical conditions, including those classified as immunocompromised, compared to BNT162b2. Findings were consistent across vaccine regimen, age group, and Delta and Omicron SARS-CoV-2 variants, as well as across patients with cancer, patients with cardiovascular, metabolic, and renal conditions, and patients receiving immunosuppressive drugs. In the absence of RCTs evaluating head-to-head VE, synthesized comparative effectiveness data provide the highest ranked evidence [126] that may guide the choice of COVID-19 vaccines for individual patients and support population-level strategies designed to prevent severe disease in high-risk populations.

## Key Summary Points

### Why carry out this study?

More than 20% of the global population has a medical condition that increases their risk of developing severe COVID-19.

Global and national health authorities recommend COVID-19 vaccination in populations with underlying medical conditions to mitigate the risk of severe illness; however, limited data are available to inform population-level policy and individual vaccination decisions.

### What was learned from the study?

To support clinical decision-making regarding COVID-19 vaccination, this systematic literature review and meta-analysis synthesized real-world effectiveness data to evaluate the comparative effectiveness of mRNA-1273 versus BNT162b2 in adults with at least one medical condition that increases the risk of developing severe COVID-19.

Vaccination with mRNA-1273 was associated with a significantly lower risk of SARS-CoV-2 infections and COVID-19–associated hospitalization and death in patients at risk of developing severe COVID-19 based on underlying medical conditions.

## Data Availability

All data produced in the present work are contained in the manuscript

## Author Contributions

XW, AP, MTB-J, MM, and EB designed and performed the systematic literature review and meta-analysis and critically evaluated the manuscript. PS, SV, RG, and MN performed the systematic literature review and critically evaluated the manuscript. AG, MGW, KZHL, PD, and NG conducted the analysis and critically evaluated the manuscript. MTB-J, NVdV, and EB conceptualized the article and provided oversight and critical evaluation of the manuscript. All authors contributed to the article and approved the submitted version.

## Acknowledgments

Writing assistance was provided by Erin McClure, PhD, and Sheri Arndt, PharmD, of ICON (Blue Bell, PA, USA) in accordance with Good Publication Practice (GPP 2022) guidelines, funded by Moderna, Inc., and under the direction of the authors.

## Data Sharing Statement

All original data generated or analyzed in this study are included in this article/as Supplementary Material. Further inquiries can be directed to the corresponding author.

