## Supplemental material for "Comparative Effectiveness of the mRNA-1273 and BNT162b2 COVID-19 Vaccines Among Adults With Underlying Medical Conditions: A Systematic Literature Review and Pairwise Meta-Analysis Using GRADE"

### Appendix – Supplemental Methods

#### *Decision Rules for Data Input into the Meta-Analysis*

- For the analysis in adults with at least one underlying medical condition, if a study reported data for more than one medical condition, the condition with the largest sample size was considered for analysis because it was possible that medical conditions were not mutually exclusive
- If there were data for more than one ≥3-dose series in a study, event numbers and sample sizes for all ≥3 dose series (homologous or heterologous) were summed for each medical condition and outcome (i.e., $(n_1+n_2+...)/(N_1+N_2+...)$). These aggregate data were used for the ≥3 doses subgroups (homologous or heterologous)
- Events reported for different categories in a study were combined as applicable (eg, various age categories for <65 years old, patients on and off tyrosine kinase inhibitors, on and off immunosuppressants, holding medication and not holding medication)
- If outcomes were reported for both 2 and ≥3 doses, ≥3 doses were considered for the primary analysis
- If a study reported outcome data for different variants but additionally reported overall data for all variants combined, the overall data were used for analysis
- If a study reported outcome data for different variants without reporting overall data for all variants combined, data for the variant with the largest sample size by vaccine arm were used for analysis
- Data for SARS-CoV-2 infections occurring ≥7 days after the last vaccination were preferentially used over <7 days in the meta-analysis if available; the earliest timepoint 7 days post-vaccination was preferred (eg, if both >7 days and >14 days were reported, >7 days was used)

- For the analysis of the severe SARS-CoV-2 infection outcome, study-defined severe infection data were used preferentially. If only hospitalization and death data were available, hospitalization data were used to derive severe infections
- If the number of events was suppressed and reported as ‘<n’ (eg, <10 or ≤5), the largest possible number of events was used for the analysis (eg, 9 or 5, respectively)
- If only vaccine effectiveness (VE) data were reported for various mutually exclusive subgroups for a particular study, and sample size was not available, the VE data for all subgroups were synthesized in a meta-analysis to obtain a risk ratio for the study, which was then used as a single input from the study in the overall meta-analysis
- If a study compared a homologous series of mRNA-1273 with a heterologous series of BNT162b2, or vice-versa, the estimated treatment effect was used in the analysis of the heterologous vaccine subgroup

#### ***Definitions of Medical Conditions***

*Clinically Extremely Vulnerable (CEV) Groups 1 and 2*

One set of base-case analyses was performed on the immunocompromised target population, which was defined as adults with a medical condition classified as CEV groups 1 or 2 [48]. Briefly, these medical conditions rendered patients severely (group 1) or moderately (group 2) immunosuppressed.

In alignment with the definitions of CEV 1 and 2, the following assumptions were made for our meta-analysis:

- Only patient populations wherein the study specified active treatment or a treatment history for cancer were categorized as CEV 1 and/or 2

- Autoimmune diseases were categorized as CEV 1 and/or 2 only if there was evidence that ≥90% of the patients were on immunosuppressive drugs
- Patients with diabetes were considered as CEV 3 irrespective of insulin treatment because most studies did not report this information

*Patients receiving immunosuppressive therapy*

Studies where ≥90% of the patient population was on immunosuppressive drugs were considered for analysis. Unless the percentage of patients on immunosuppressive therapies was specified, all patients with solid organ transplant, hematologic malignancies, and chronic kidney disease with hemodialysis were assumed to be receiving immunosuppressive drugs. The following drugs were considered to be immunosuppressants:

- Glucocorticoids
- Cytostatics (eg, methotrexate, azathioprine, mercaptopurine)
- Antibodies (eg, monoclonal antibodies, alemtuzumab, muromonab, rituximab, belatacept)
- Drugs acting on immunophilins (eg, cyclosporine, tacrolimus, sirolimus, everolimus, zotarolimus)
- Other drugs (eg, interferons, opioids, tumor necrosis factor-binding proteins, mycophenolate, small biological agents)

*Cancers*

Study populations with hematologic malignancies, solid tumors, mixed cancers, or unspecified cancer types were included in this subgroup.

*Cardiovascular-metabolic-renal disorders*

Study populations with cardiovascular disease (including hypertension), type 1 or type 2 diabetes, obesity, and chronic kidney disease with or without hemodialysis were included in this subgroup.

*Mixed conditions*

Mixed conditions includes a combination of various medical conditions including hypertension, diabetes, chronic lung disease, immunodeficiencies, heart disease, renal disease, liver disease, immunosuppression. It additionally includes a mixed population of organ and stem cell transplant.

*Immunocompromised*

Populations defined as “immunocompromised” in the study and/or receiving immunosuppressive therapies.

**Table S1** Databases and search strings used for the systematic literature review

| **Embase** | |
|---|---|
| **No.** | **Search terms** |
| 1 | exp coronavirus disease 2019/ |
| 2 | exp Coronavirus infection/ or exp Coronavirus/ or exp Coronavirinae/ |
| 3 | exp Coronaviridae infection/ or exp Coronaviridae/ |
| 4 | exp SARS coronavirus/ or exp SARS-related coronavirus/ |
| 5 | exp Severe acute respiratory syndrome coronavirus 2/ |
| 6 | (sars cov2 or sars cov 2 or sarscov2 or sarscov 2 or sars-cov-2 or sarscov-2).ti,ab,tw. |
| 7 | Severe acute respiratory syndrome coronavirus.ti,ab,tw. |
| 8 | or/1-7 |
| 9 | exp elasomeran/ |
| 10 | exp imelasomeran/ or exp elasomeran plus imelasomeran/ |
| 11 | exp davesomeran/ or exp davesomeran plus elasomeran/ |
| 12 | (Moderna or Spikevax or elasomeran or imelasomeran or davesomeran or andusomeran or mRNA-1273 or mRNA1273 or mRNA 1273 or messenger* or messenger RNA-1273 or messenger RNA 1273 or messenger RNA1273).ti,ab,tw,kf,mp. |
| 13 | or/9-12 |
| 14 | exp bnt 162 vaccine/ |
| 15 | exp tozinameran/ |
| 16 | exp riltozinameran/ or exp riltozinameran plus tozinameran/ |
| 17 | exp famtozinameran/ or exp famtozinameran plus tozinameran/ |
| 18 | (Pfizer or PfizerBioNTech or Pfizer-BioNTech or pfizer* or 'BioNTech' or BioNTech-Pfizer or COMIRNATY or Tozinameran or riltozinameran or famtozinameran or raxtozinameran or BNT162b2 or BNT*).ti,ab,tw,kf,mp. |

| | |
|---|---|
| 19 | or/14-18 |
| 20 | 13 and 19 |
| 21 | 8 and 20 |
| 22 | exp comparative effectiveness/ or exp clinical effectiveness/ |
| 23 | (efficacy or effectiveness or infection or infected or infections).ti,ab,tw. |
| 24 | exp hospital mortality/ or exp mortality/ |
| 25 | (hospitali$ed or hospitali$ation$1 or admission$1).ti,ab,tw. |
| 26 | exp death/ |
| 27 | exp breakthrough infection/ or exp infection fatality rate/ or exp infection rate/ or exp infection/ or exp asymptomatic infection/ |
| 28 | exp reinfection/ |
| 29 | exp hospital admission/ or exp hospitalization/ |
| 30 | (mortality or death$1).ti,ab,tw. |
| 31 | (symptomatic or asymptomatic or severe infection or laboratory confirmed or laboratory-confirmed or pneumonia or respiratory failure or test positive or test-positive or re-infection or re infection or reinfection or breakthrough infection or positiv* or infect*).ti,ab,tw. |
| 32 | or/22-31 |
| 33 | 21 and 32 |
| 34 | exp autoimmune disease/ or exp steroid/ or exp rheumatic disease/ or exp autoimmunity/ or exp inflammatory disease/ |
| 35 | exp psoriasis/ or exp inflammatory bowel disease/ or exp multiple sclerosis/ or exp systemic lupus erythematous/ or exp ankylosing spondylitis/ |
| 36 | (steroid* or rheumatic or rheumatoid or psoriasis or psoriatic or lupus or SLE or ulcerative colitis or crohn$2 or Inflammatory bowel disease or IBD or Addison$2 or Grave$2 or Hashimoto$2 or Myasthenia gravis or celiac or multiple sclerosis or pernicious or autoimmune or ankylosing spondylitis or immune mediated inflammatory disease or immune-mediated or immune mediated or inflammatory disease* or IMID).ti,ab,tw. |

| | |
|---|---|
| 37 | or/34-36 |
| 38 | exp solid tumor/ |
| 39 | exp malignant neoplasm/ or exp cancer/ or exp malignancy/ |
| 40 | exp antineoplastic agent/ or exp neoadjuvant chemotherapy/ or *chemotherapy/ or exp cancer combination chemotherapy/ or *cancer chemotherapy/ |
| 41 | (solid tumor or cancer or carcinoma or malignan* or neoplasm or chemotherap* or antineoplastic or anti-neoplastic or cytotoxic therapy or anti-cancer or anticancer).ti,ab,tw. |
| 42 | or/38-41 |
| 43 | exp graft recipient/ or exp transplantation/ or exp organ transplantation/ or exp transplant/ |
| 44 | (transplant* or solid organ or solid organ transplant*).ti,ab,tw. |
| 45 | or/43-44 |
| 46 | exp hematologic malignancy/ |
| 47 | exp stem cell/ or exp hematopoietic stem cell transplantation/ |
| 48 | (transplant* or stem cell or SCT or HSCT or BMT or bone marrow).ti,ab,tw. |
| 49 | (inherited red blood cell disorders or haemoglobinopath* or hemoglobinopath* or hemolytic or haemolytic or hemato* or haemato*).ti,ab. |
| 50 | or/46-49 |
| 51 | exp chronic kidney failure/ or exp chronic kidney disease/ or exp nephrotic syndrome/ |
| 52 | (chronic kidney or CKD or chronic nephrotic or hemodialysis or haemodialysis).ti,ab,tw. |
| 53 | or/51-52 |
| 54 | exp diabetes mellitus/ or exp non insulin dependent diabetes mellitus/ or exp insulin dependent diabetes mellitus/ |

| | |
|---|---|
| 55 | (diabet* or T1DM or T2DM or TIDM or TIIDM or IDDM or NIDDM or type 1 diabetes or type I diabetes or type 2 diabetes or type II diabetes or insulin or non-insulin).ti,ab,tw. |
| 56 | 54 or 55 |
| 57 | exp cardiovascular disease/ |
| 58 | exp ischemic heart disease/ or exp coronary artery disease/ or exp angina pectoris/ or exp heart rehabilitation/ |
| 59 | exp hypertension/ or exp myocarditis/ |
| 60 | (heart or cardiac or coronary or cardio* or CHD or high blood pressure or hypertension or myocard* or cardiovasc* or cardiomyopath* or CAD or heart failure or arterial or arter* or aortic).ti,ab,tw. |
| 61 | or/57-60 |
| 62 | exp chronic liver disease/ or exp liver cirrhosis/ or exp chronic hepatitis/ |
| 63 | exp non alcoholic fatty liver/ or exp alcoholic liver disease/ or exp autoimmune hepatitis/ |
| 64 | exp liver transplantation/ |
| 65 | (chronic liver or CLD or cirrho* or chronic hepatitis or non-alcoholic fatty liver or NAFLD or non alcoholic fatty liver or alcoholic liver or autoimmune hepatitis or chronic hepatic).ti,ab,tw. |
| 66 | or/62-65 |
| 67 | exp chronic lung disease/ or exp lung embolism/ or exp chronic pulmonary aspergillosis/ or exp pulmonary hypertension/ |
| 68 | exp chronic obstructive lung disease/ or exp asthma/ or exp lung dysplasia/ |
| 69 | exp lung emphysema/ or exp emphysema/ or exp interstitial lung disease/ or exp fibrosing alveolitis/ or exp cystic fibrosis/ |
| 70 | (asthma* or bronchiectasis or bronchopulmonary dysplasia or chronic lung or chronic pulmonary or chronic obstructive or COPD or emphysema or interstitial lung disease or idiopathic pulmonary fibrosis or pulmonary embolism or pulmonary hypertension or cystic fibrosis).ti,ab,tw. |
| 71 | or/67-70 |

| | |
|---|---|
| 72 | exp obesity/ |
| 73 | (overweight or obes*).ti,ab,tw. |
| 74 | or/72-73 |
| 75 | exp dementia/ or exp Alzheimer disease/ or exp brain disease/ or exp degenerative disease/ or exp brain degeneration/ or exp neurologic disease/ or exp amyotrophic lateral sclerosis/ |
| 76 | (dementia or Alzheimer$2 or degenerative brain or degenerative mental or neurolog* or neurodegen* or amyotrophic lateral sclerosis or ALS or Parkinson$2 or epilepsy or Guillain$2).ti,ab,tw. |
| 77 | or/75-76 |
| 78 | exp cerebrovascular accident/ or exp cerebrovascular disease/ or exp cerebrovascular malformation/ |
| 79 | (stroke or cerebrovascular or cerebral or ischemic or ischaemic or ischem* or ischaem* or carotid or aneurysm* or arteriovenous or arterio-venous).ti,ab,kw. |
| 80 | or/78-79 |
| 81 | exp immunocompromised patient/ or exp immune deficiency/ or exp immunosuppressive treatment/ |
| 82 | (immunocompr* or immunodef* or immunosupp* or immuno-compromised or immuno-deficiency or immuno-suppression or immunomod*).ti,ab. |
| 83 | or/81-82 |
| 84 | exp *comorbidity/ or exp *long term care/ or exp *chronic disease/ |
| 85 | (comorbid* or risk condition or risk disorder or at-risk or at risk or high risk or high-risk or risk disease or risk comorbidity or risk disability or long term condition or long-term condition or underlying* or pre-existing* or preexisting* or medical condition).ti,ab,kw. |
| 86 | or/84-85 |
| 87 | 37 or 42 or 45 or 50 or 53 or 56 or 61 or 66 or 71 or 74 or 77 or 80 or 83 or 86 |
| 88 | 33 and 87 |
| 89 | Clinical study/ |

| | |
|---|---|
| 90 | exp case control study/ |
| 91 | Family study/ |
| 92 | Longitudinal study/ |
| 93 | Retrospective study/ |
| 94 | Prospective study/ |
| 95 | Randomized controlled trials/ |
| 96 | 94 not 95 |
| 97 | Cohort analysis/ |
| 98 | (Cohort adj (study or studies)).mp. |
| 99 | (Case control adj (study or studies)).tw. |
| 100 | (follow up adj (study or studies)).tw. |
| 101 | (observational adj (study or studies)).tw. |
| 102 | (epidemiologic$ adj (study or studies)).tw. |
| 103 | (cross sectional adj (study or studies)).tw. |
| 104 | or/89-93,96-103 |
| 105 | 88 and 104 |
| 106 | Clinical Trial/ |
| 107 | Randomized Controlled Trial/ |
| 108 | controlled clinical trial/ |
| 109 | multicenter study/ |
| 110 | Phase 3 clinical trial/ |
| 111 | Phase 4 clinical trial/ |
| 112 | RANDOMIZATION/ |
| 113 | Single Blind Procedure/ |

| | |
|---|---|
| 114 | Double Blind Procedure/ |
| 115 | Crossover Procedure/ |
| 116 | PLACEBO/ |
| 117 | randomi?ed controlled trial$.tw. |
| 118 | rct.tw. |
| 119 | (random$ adj2 allocat$).tw. |
| 120 | single blind$.tw. |
| 121 | double blind$.tw. |
| 122 | ((treble or triple) adj blind$).tw. |
| 123 | placebo$.tw. |
| 124 | Prospective Study/ |
| 125 | or/106-124 |
| 126 | Case Study/ |
| 127 | case report.tw. |
| 128 | or/126-127 |
| 129 | 125 not 128 |
| 130 | 88 and 129 |
| 131 | 105 or 130 |
| 132 | limit 131 to (human and english language) |
| 133 | limit 132 to yr="2019 -Current" |

| **Ovid MELINE®** | |
|---|---|
| **No.** | **Search terms** |
| 1 | exp coronavirus disease 2019/ |
| 2 | exp Coronavirus infection/ or exp Coronavirus/ or exp Coronavirinae/ |

| | |
|---|---|
| 3 | exp Coronaviridae infection/ or exp Coronaviridae/ |
| 4 | exp SARS coronavirus/ or exp SARS-related coronavirus/ |
| 5 | exp Severe acute respiratory syndrome coronavirus 2/ |
| 6 | (sars cov2 or sars cov 2 or sarscov2 or sarscov 2 or sars-cov-2 or sarscov-2).ti,ab,tw. |
| 7 | Severe acute respiratory syndrome coronavirus.ti,ab,tw. |
| 8 | or/1-7 |
| 9 | RNA vaccine/ or mRNA vaccine$1.ti,ab,tw. |
| 10 | exp elasomeran/ |
| 11 | (Moderna or Spikevax$1 or elasomeran or imelasomeran or davesomeran or andusomeran or mRNA-1273 or mRNA1273 or mRNA 1273).ti,ab,tw,kf,mp. |
| 12 | 10 or 11 |
| 13 | exp tozinameran/ |
| 14 | (Pfizer or PfizerBioNTech or Pfizer-BioNTech or pfizer* or 'BioNTech' or BioNTech-Pfizer or COMIRNATY or Tozinameran or BNT162b2 or BNT* or riltozinameran or famtozinameran or raxtozinameran).ti,ab,tw,kf,mp. |
| 15 | 13 or 14 |
| 16 | 9 or (12 and 15) |
| 17 | 8 and 16 |
| 18 | exp comparative effectiveness/ or exp clinical effectiveness/ |
| 19 | (efficacy or effectiveness or infection or infected or infections).ti,ab,tw. |
| 20 | exp hospital mortality/ or exp mortality/ |
| 21 | (hospitali$ed or hospitali$ation$1 or admission$1).ti,ab,tw. |
| 22 | exp death/ |
| 23 | exp breakthrough infection/ or exp infection fatality rate/ or exp infection rate/ or exp infection/ or exp asymptomatic infection/ |

| | |
|---|---|
| 24 | exp reinfection/ |
| 25 | exp hospital admission/ or exp hospitalization/ |
| 26 | (mortality or death$1).ti,ab,tw. |
| 27 | (symptomatic or asymptomatic or severe infection or laboratory confirmed or laboratory-confirmed or pneumonia or respiratory failure or test positive or test-positive or re-infection or re infection or reinfection or breakthrough infection or positiv* or infect*).ti,ab,tw. |
| 28 | or/18-27 |
| 29 | 17 and 28 |
| 30 | exp autoimmune disease/ or exp steroid/ or exp rheumatic disease/ or exp autoimmunity/ |
| 31 | exp psoriasis/ or exp inflammatory bowel disease/ or exp multiple sclerosis/ or exp systemic lupus erythematous/ |
| 32 | (steroid* or rheumatic or rheumatoid or psoriasis or psoriatic or lupus or SLE or ulcerative colitis or crohn$2 or Inflammatory bowel disease or IBD or Addison$2 or Grave$2 or Hashimoto$2 or Myasthenia gravis or celiac or multiple sclerosis or pernicious or autoimmune or ankylosing spondylitis or immune mediated inflammatory disease or immune-mediated or immune mediated or inflammatory disease* or IMID).ti,ab,tw. |
| 33 | or/30-32 |
| 34 | exp Neoplasms/ |
| 35 | exp malignant neoplasm/ or exp cancer/ or exp malignancy/ |
| 36 | exp antineoplastic agent/ or exp neoadjuvant chemotherapy/ or *chemotherapy/ or exp cancer combination chemotherapy/ or *cancer chemotherapy/ |
| 37 | (solid tumor or cancer or carcinoma or malignan* or neoplasm or chemotherap* or antineoplastic or anti-neoplastic or cytotoxic therapy or anti-cancer or anticancer).ti,ab,tw. |
| 38 | or/34-37 |
| 39 | exp graft recipient/ or exp transplantation/ or exp organ transplantation/ or exp transplant/ |

| | |
|---|---|
| 40 | (transplant* or solid organ or solid organ transplant*).ti,ab,tw. |
| 41 | or/39-40 |
| 42 | exp hematologic malignancy/ |
| 43 | exp stem cell/ or exp hematopoietic stem cell transplantation/ |
| 44 | (transplant* or stem cell or SCT or HSCT or BMT or bone marrow).ti,ab,tw. |
| 45 | (inherited red blood cell disorders or haemoglobinopath* or hemoglobinopath* or hemolytic or haemolytic or hemato* or haemato*).ti,ab. |
| 46 | or/42-45 |
| 47 | exp chronic kidney failure/ or exp chronic kidney disease/ or exp nephrotic syndrome/ |
| 48 | (chronic kidney or CKD or chronic nephrotic or hemodialysis or haemodialysis).ti,ab,tw. |
| 49 | or/47-48 |
| 50 | exp diabetes mellitus/ or exp non insulin dependent diabetes mellitus/ or exp insulin dependent diabetes mellitus/ |
| 51 | (diabet* or T1DM or T2DM or TIDM or TIIDM or IDDM or NIDDM or type 1 diabetes or type I diabetes or type 2 diabetes or type II diabetes or insulin or non-insulin).ti,ab,tw. |
| 52 | 50 or 51 |
| 53 | exp cardiovascular disease/ |
| 54 | exp ischemic heart disease/ or exp coronary artery disease/ or exp angina pectoris/ or exp heart rehabilitation/ |
| 55 | exp hypertension/ or exp myocarditis/ |
| 56 | (heart or cardiac or coronary or cardio* or CHD or high blood pressure or hypertension or myocard* or cardiovasc*).ti,ab,tw. |
| 57 | or/53-56 |
| 58 | exp chronic liver disease/ or exp liver cirrhosis/ or exp chronic hepatitis/ |

| | |
|---|---|
| 59 | exp non alcoholic fatty liver/ or exp alcoholic liver disease/ or exp autoimmune hepatitis/ |
| 60 | exp liver transplantation/ |
| 61 | (chronic liver or CLD or cirrho* or chronic hepatitis or non-alcoholic fatty liver disease or NAFLD or alcoholic liver disease or autoimmune hepatitis or chronic hepatic).ti,ab,tw. |
| 62 | or/58-61 |
| 63 | exp chronic lung disease/ or exp lung embolism/ or exp chronic pulmonary aspergillosis/ or exp pulmonary hypertension/ |
| 64 | exp chronic obstructive lung disease/ or exp asthma/ or exp lung dysplasia/ |
| 65 | exp lung emphysema/ or exp emphysema/ or exp interstitial lung disease/ or exp fibrosing alveolitis/ or exp cystic fibrosis/ |
| 66 | (asthma* or bronchiectasis or bronchopulmonary dysplasia or chronic lung or chronic pulmonary or chronic obstructive or COPD or emphysema or interstitial lung disease or idiopathic pulmonary fibrosis or pulmonary embolism or pulmonary hypertension or cystic fibrosis).ti,ab,tw. |
| 67 | or/63-66 |
| 68 | exp obesity/ |
| 69 | (overweight or obes*).ti,ab,tw. |
| 70 | or/68-69 |
| 71 | exp dementia/ or exp Alzheimer disease/ or exp brain disease/ or exp degenerative disease/ or exp brain degeneration/ or exp neurologic disease/ or exp amyotrophic lateral sclerosis/ |
| 72 | (dementia or Alzheimer$2 or degenerative brain or degenerative mental or neurolog* or neurodegen* or amyotrophic lateral sclerosis or ALS or Parkinson$2 or epilepsy or Guillain$2).ti,ab,tw. |
| 73 | or/71-72 |
| 74 | exp cerebrovascular disorder/ or stroke/ |
| 75 | (cerebrovascular or stroke or cerebral or carotid or ischemia or ischaemia or aneurysm* or arteriovenous or arterio-venous).ti,ab,tw. |

| | |
|---|---|
| 76 | or/74-75 |
| 77 | exp immunocompromised patient/ or exp immune deficiency/ or exp immunosuppressive treatment/ |
| 78 | (immunocompr* or immunodef* or immunosupp* or immuno-compromised or immuno-deficiency or immuno-suppression or immunomod*).ti,ab,kw. |
| 79 | or/77-78 |
| 80 | exp *comorbidity/ or exp *long term care/ or exp *chronic disease/ |
| 81 | (comorbid* or risk condition or risk disorder or at-risk or at risk or high risk or high-risk or risk disease or risk comorbidity or risk disability or long term condition or long-term condition or underlying* or pre-existing* or preexisting* or medical condition).ti,ab,kw. |
| 82 | or/80-81 |
| 83 | 33 or 38 or 41 or 46 or 49 or 52 or 57 or 62 or 67 or 70 or 73 or 76 or 79 or 82 |
| 84 | 29 and 83 |
| 85 | Epidemiologic studies/ |
| 86 | exp case control studies/ |
| 87 | exp cohort studies/ |
| 88 | Case control.tw. |
| 89 | (cohort adj (study or studies)).tw. |
| 90 | Cohort analy$.tw. |
| 91 | (Follow up adj (study or studies)).tw. |
| 92 | (observational adj (study or studies)).tw. |
| 93 | Longitudinal.tw. |
| 94 | Retrospective.tw. |
| 95 | Cross sectional.tw. |
| 96 | Cross-sectional studies/ |

| | |
|---|---|
| 97 | or/85-96 |
| 98 | 84 and 97 |
| 99 | Randomized Controlled Trials as Topic/ |
| 100 | randomized controlled trial/ |
| 101 | Random Allocation/ |
| 102 | Double Blind Method/ |
| 103 | Single Blind Method/ |
| 104 | clinical trial/ |
| 105 | clinical trial, phase i.pt. |
| 106 | clinical trial, phase ii.pt. |
| 107 | clinical trial, phase iii.pt. |
| 108 | clinical trial, phase iv.pt. |
| 109 | controlled clinical trial.pt. |
| 110 | randomized controlled trial.pt. |
| 111 | multicenter study.pt. |
| 112 | clinical trial.pt. |
| 113 | exp Clinical Trials as topic/ |
| 114 | or/99-113 |
| 115 | (clinical adj trial$).tw. |
| 116 | ((singl$ or doubl$ or treb$ or tripl$) adj (blind$3 or mask$3)).tw. |
| 117 | PLACEBOS/ |
| 118 | placebo$.tw. |
| 119 | randomly allocated.tw. |
| 120 | (allocated adj2 random$).tw. |

| | |
|---|---|
| 121 | or/115-120 |
| 122 | 114 or 121 |
| 123 | case report.tw. |
| 124 | letter/ |
| 125 | historical article/ |
| 126 | or/123-125 |
| 127 | 122 not 126 |
| 128 | 84 and 127 |
| 129 | 98 or 128 |
| 130 | limit 129 to (english language and humans) |
| 131 | limit 130 to yr="2019 -Current" |
| **EBM Reviews – Cochrane Central Register of Controlled Trials** | |
| **No.** | **Search terms** |
| 1 | exp coronavirus disease 2019/ |
| 2 | exp Coronavirus infection/ or exp Coronavirus/ or exp Coronavirinae/ |
| 3 | exp Coronaviridae infection/ or exp Coronaviridae/ |
| 4 | exp Severe acute respiratory syndrome coronavirus 2/ |
| 5 | (sars cov2 or sars cov 2 or sarscov2 or sarscov 2 or sars-cov-2 or sarscov-2).ti,ab,tw. |
| 6 | Severe acute respiratory syndrome coronavirus.ti,ab,tw. |
| 7 | or/1-6 |
| 8 | RNA vaccine/ |
| 9 | mRNA vaccine$1.ti,ab,tw. |
| 10 | (Moderna or Spikevax or elasomeran or imelasomeran or davesomeran or andusomeran or mRNA-1273 or mRNA1273 or mRNA 1273).ti,ab,tw,mp. |

| | |
|---|---|
| 11 | (Pfizer or PfizerBioNTech or Pfizer-BioNTech or pfizer* or 'BioNTech' or BioNTech-Pfizer or COMIRNATY or Tozinameran or BNT162b2 or BNT* or riltozinameran or famtozinameran or raxtozinameran).ti,ab,tw,mp. |
| 12 | 8 or 9 or (10 and 11) |
| 13 | 7 and 12 |

**EBM Reviews – Cochrane Database of Systematic Reviews**

| **No.** | **Search terms** |
|---|---|
| 1 | exp coronavirus disease 2019/ |
| 2 | exp Coronavirus infection/ or exp Coronavirus/ or exp Coronavirinae/ |
| 3 | exp Coronaviridae infection/ or exp Coronaviridae/ |
| 4 | exp Severe acute respiratory syndrome coronavirus 2/ |
| 5 | (sars cov2 or sars cov 2 or sarscov2 or sarscov 2 or sars-cov-2 or sarscov-2).ti,ab,tw. |
| 6 | Severe acute respiratory syndrome coronavirus.ti,ab,tw. |
| 7 | or/1-6 |
| 8 | RNA vaccine/ |
| 9 | mRNA vaccine$1.ti,ab,tw. |
| 10 | (Moderna or Spikevax or elasomeran or imelasomeran or davesomeran or andusomeran or mRNA-1273 or mRNA1273 or mRNA 1273).ti,ab,tw,mp. |
| 11 | (Pfizer or PfizerBioNTech or Pfizer-BioNTech or pfizer* or 'BioNTech' or BioNTech-Pfizer or COMIRNATY or Tozinameran or BNT162b2 or BNT* or riltozinameran or famtozinameran or raxtozinameran).ti,ab,tw,mp. |
| 12 | 8 or 9 or (10 and 11) |
| 13 | 7 and 12 |

**Table S2** Summary of population, intervention, comparison, and outcomes used in systematic literature review

| | | |
|---|---|---|
| **Research question** | Following primary series vaccination and additional vaccine doses, is mRNA-1273 more effective than BNT162b2 at preventing SARS-CoV-2 infections and COVID-19-related hospitalizations and deaths in people with underlying medical conditions? | |
| | **Include** | **Exclude** |
| **Population** | Adults ≥18 years old with the following medical conditions:<br>• Autoimmune disease<br>• Solid tumor<br>• Solid organ transplant<br>• Hematologic malignancy<br>• Chronic kidney disease with and without hemodialysis<br>• Type 1 and type 2 diabetes<br>• Cardiovascular disease (hypertension, heart failure, coronary artery disease, cardiomyopathies)<br>• Cerebrovascular disease<br>• Chronic liver disease (cirrhosis, non-alcoholic fatty liver disease, alcoholic liver disease, autoimmune hepatitis)<br>• Neurologic condition (dementia, Alzheimer’s disease, ALS, Parkinson’s disease)<br>• Chronic respiratory conditions (asthma, bronchiectasis, COPD, interstitial lung disease, pulmonary embolism, pulmonary hypertension)<br>• Obesity<br><br>Adults described as “immunocompromised” or with | • Pregnant women<br>• Children |

| | | |
|---|---|---|
| | primary immunodeficiency syndrome<br><br>Adults with mixed cancers or "cancer"<br><br>Studies that included a combination of any of the above-mentioned populations with data reported as overall effectiveness, not by specific medical condition<br><br>Studies on the general population with ≥90% of the population with ≥1 medical condition | |
| **Interventions/ Comparators** | mRNA COVID-19 vaccines, including strain-updated versions<br><br>• Moderna COVID-19 vaccine (mRNA-1273, SPIKEVAX, elasomeran, imelasomeran, davesomeran, andusomeran)<br>• Pfizer/BioNTech COVID-19 vaccine (BNT162b2, COMIRNATY, tozinameran, riltozinameran, famtozinameran, raxtozinameran)<br><br>**Dosing regimens**<br><br>• Homologous 2-dose series<br>• Homologous and heterologous series of ≥3 doses with Moderna or Pfizer/BioNTech vaccines as the last dose | • Heterologous 2-dose series<br>• Moderna COVID-19 vaccine as the last dose vs other COVID-19 vaccines as the last dose (eg, Janssen, AstraZeneca)<br>• Pfizer/BioNTech COVID-19 vaccine as the last dose vs other COVID-19 vaccines as the last dose<br>• Moderna and Pfizer/BioNTech COVID-19 vaccines as the primary series vs other COVID-19 vaccines as the primary series |
| **Outcomes** | • SARS-CoV-2 infection (positive test or symptomatic)<br>• Symptomatic laboratory-confirmed SARS-CoV-2 infection<br>• Severe SARS-CoV-2 infection<br>• Breakthrough SARS-CoV-2 infection | Studies with only safety and immunogenicity outcomes |

| | | |
|---|---|---|
| | • SARS-CoV-2 re-infection<br>• Hospitalization due to COVID-19 (eg, ICU, ER, or ventilation)<br>• Death due to COVID-19 | |
| **Study design** | • Randomized and nonrandomized clinical trials<br>• Observational studies | • Study protocols<br>• Economic models<br>• Case reports<br>• Review articles |
| **Other limits** | • Any publication type (including letter, commentary, abstract, full text, poster, case series)<br>• English language | |

ALS, amyotrophic lateral sclerosis; COPD, chronic obstructive pulmonary disease; ER, emergency room; ICU, intensive care unit.

**Table S3** Results from risk of bias assessment based on the Newcastle-Ottawa Scale[a]

| Cohort Studies | | | | | | | | | |
|---|---|---|---|---|---|---|---|---|---|
| | **Patient selection** | | | | **Comparability of cohorts[f]** | **Outcome** | | | **Total score** |
| **Author, year** | **Representativeness of exposed cohort[b]** | **Selection of nonexposed cohort[c]** | **Ascertainment of exposure[d]** | **Outcome not present at baseline[e]** | | **Assessment of outcome[g]** | **Sufficient follow-up duration[h]** | **Adequate follow-up[i]** | |
| Aslam, 2021 [72] | 1 star | 1 star | 1 star | 1 star | 0 star | 1 star | 1 star | 1 star | 7 star |
| Bello-Chavolla, 2023 [103] | 0 star | 1 star | 1 star | 1 star | 1 star | 1 star | 1 star | 1 star | 7 star |
| Boekel, 2022 [120] | 1 star | 1 star | 1 star | 0 star | 0 star | 1 star | 1 star | 1 star | 6 star |
| Bonazzetti, 2023 [111] | 1 star | 1 star | 1 star | 1 star | 1 star | 1 star | 1 star | 1 star | 8 star |
| Capuano, 2023 [112] | 1 star | 1 star | 1 star | 0 star | 0 star | 1 star | 1 star | 1 star | 6 star |
| Chen, 2023 [124] | 1 star | 1 star | 1 star | 0 star | 0 star | 1 star | 0 star | 0 star | 4 star |
| Cona, 2023 [113] | 1 star | 1 star | 1 star | 1 star | 0 star | 1 star | 1 star | 1 star | 7 star |
| Cook, 2023 [74] | 1 star | 1 star | 1 star | 0 star | 1 star | 1 star | 1 star | 1 star | 7 star |
| Dimitrov, 2023 [116] | 1 star | 1 star | 1 star | 0 star | 0 star | 1 star | 1 star | 1 star | 6 star |
| Egri, 2023 [104] | 0 star | 1 star | 1 star | 1 star | 0 star | 1 star | 1 star | 1 star | 6 star |
| Embi, 2021 [76] | 1 star | 1 star | 1 star | 1 star | 1 star | 1 star | 0 star | 0 star | 6 star |
| Embi, 2023 [77] | 1 star | 1 star | 1 star | 1 star | 0 star | 1 star | 1 star | 1 star | 7 star |
| Figueiredo, 2021 [78] | 1 star | 0 star | 1 star | 0 star | 1 star | 1 star | 1 star | 0 star | 5 star |
| Fu, 2023 [79] | 1 star | 1 star | 1 star | 0 star | 1 star | 1 star | 1 star | 1 star | 7 star |

| | | | | | | | | | |
|---|---|---|---|---|---|---|---|---|---|
| Heinzl, 2022 [118] | 1 star | 1 star | 1 star | 1 star | 1 star | 1 star | 0 star | 1 star | 7 star |
| Hernandez, 2022 [61] | 1 star | 0 star | 1 star | 1 star | 0 star | 1 star | 1 star | 1 star | 6 star |
| Holroyd, 2022 [80] | 0 star | 1 star | 1 star | 1 star | 1 star | 1 star | 1 star | 1 star | 7 star |
| John, 2022 [82] | 1 star | 1 star | 1 star | 1 star | 1 star | 1 star | 1 star | 0 star | 7 star |
| Kelly, 2022 [84] | 1 star | 1 star | 1 star | 1 star | 1 star | 1 star | 1 star | 1 star | 8 star |
| Kelly, 2023 [83] | 1 star | 1 star | 1 star | 1 star | 0 star | 1 star | 1 star | 1 star | 7 star |
| Khan, 2021 [85] | 1 star | 1 star | 1 star | 1 star | 1 star | 1 star | 1 star | 1 star | 8 star |
| Kopel, 2024 [67] | 1 star | 1 star | 1 star | 1 star | 1 star | 1 star | 1 star | 1 star | 8 star |
| Kshirsagar, 2022 [86] | 1 star | 1 star | 1 star | 0 star | 0 star | 1 star | 1 star | 1 star | 6 star |
| Liew, 2022 [87] | 1 star | 0 star | 1 star | 0 star | 0 star | 1 star | 1 star | 1 star | 5 star |
| Magnusson, 2021 [121] | 1 star | 1 star | 1 star | 0 star | 0 star | 1 star | 1 star | 1 star | 6 star |
| Malinis, 2021 [88] | 1 star | 0 star | 1 star | 1 star | 0 star | 1 star | 1 star | 1 star | 6 star |
| Manley, 2023 [62] | 1 star | 1 star | 1 star | 0 star | 1 star | 1 star | 1 star | 0 star | 6 star |
| Marinaki, 2022 [119] | 1 star | 1 star | 0 star | 0 star | 0 star | 1 star | 1 star | 0 star | 4 star |
| Mazuecos, 2022 [105] | 1 star | 1 star | 1 star | 1 star | 1 star | 1 star | 1 star | 1 star | 8 star |
| Miao, 2023 [89] | 1 star | 1 star | 1 star | 1 star | 1 star | 1 star | 0 star | 0 star | 6 star |
| Motwani, 2023 [63] | 1 star | 0 star | 1 star | 1 star | 0 star | 1 star | 1 star | 1 star | 6 star |
| Mues, 2022 [64] | 1 star | 1 star | 1 star | 1 star | 2 star | 1 star | 1 star | 1 star | 9 star |

| | | | | | | | | | |
|---|---|---|---|---|---|---|---|---|---|
| Odriozola, 2022 [106] | 1 star | 0 star | 1 star | 0 star | 0 star | 1 star | 1 star | 0 star | 4 star |
| Parsons, 2023 [91] | 1 star | 0 star | 1 star | 1 star | 1 star | 1 star | 1 star | 1 star | 7 star |
| Patel, 2023 [92] | 1 star | 1 star | 1 star | 0 star | 1 star | 1 star | 1 star | 1 star | 7 star |
| Pinana, 2022 [107] | 1 star | 1 star | 1 star | 1 star | 0 star | 1 star | 1 star | 1 star | 7 star |
| Pinana, 2023a [109] | 1 star | 1 star | 1 star | 1 star | 1 star | 1 star | 1 star | 1 star | 8 star |
| Pinana, 2023b [108] | 1 star | 1 star | 1 star | 1 star | 1 star | 1 star | 1 star | 1 star | 8 star |
| Pino, 2022 [114] | 1 star | 1 star | 1 star | 1 star | 1 star | 1 star | 1 star | 1 star | 8 star |
| Quiroga, 2022 [68] | 1 star | 1 star | 1 star | 0 star | 1 star | 1 star | 1 star | 1 star | 7 star |
| Radcliffe, 2022 [100] | 1 star | 0 star | 1 star | 1 star | 0 star | 1 star | 0 star | 0 star | 4 star |
| Risk, 2022 [93] | 1 star | 1 star | 1 star | 1 star | 1 star | 1 star | 1 star | 1 star | 8 star |
| Rodriguez-Mora, 2023 [110] | 1 star | 1 star | 1 star | 1 star | 0 star | 1 star | 1 star | 1 star | 7 star |
| Rooney, 2022 [94] | 1 star | 1 star | 0 star | 1 star | 0 star | 1 star | 1 star | 1 star | 6 star |
| Shen, 2022 [95] | 1 star | 1 star | 1 star | 1 star | 0 star | 1 star | 1 star | 1 star | 7 star |
| Sibbel, 2022 [96] | 1 star | 1 star | 1 star | 1 star | 1 star | 1 star | 1 star | 1 star | 8 star |
| Song, 2022 [70] | 1 star | 1 star | 1 star | 0 star | 1 star | 1 star | 1 star | 0 star | 6 star |
| Sormani, 2022 [115] | 1 star | 0 star | 1 star | 1 star | 1 star | 1 star | 1 star | 1 star | 7 star |
| Sun, 2024 [66] | 1 star | 1 star | 1 star | 1 star | 1 star | 1 star | 1 star | 1 star | 8 star |
| Valkov, 2023 [117] | 1 star | 1 star | 1 star | 0 star | 1 star | 1 star | 0 star | 0 star | 5 star |
| Wang, 2022 [98] | 1 star | 1 star | 1 star | 1 star | 1 star | 1 star | 0 star | 0 star | 6 star |

| | | | | | | | | | |
|---|---|---|---|---|---|---|---|---|---|
| Wing, 2023 [102] | 1 star | 0 star | 1 star | 1 star | 1 star | 1 star | 1 star | 1 star | 7 star |
| Yeo, 2022 [123] | 0 star | 1 star | 1 star | 1 star | 1 star | 1 star | 1 star | 1 star | 7 star |
| Yetmar, 2022 [99] | 0 star | 0 star | 1 star | 0 star | 0 star | 1 star | 1 star | 0 star | 3 star |
| **Case Control Studies** | | | | | | | | | |
| **Author, year** | **Selection** | | | | **Comparability[n]** | **Exposure** | | | **Total score** |
| | **Is the case definition adequate?[j]** | **Representativeness of the cases[k]** | **Selection of controls[l]** | **Definition of controls[m]** | | **Ascertainment of exposure[o]** | **Same method of ascertainment for cases and controls[p]** | **Non-response rate[q]** | |
| Alkadi, 2022 [69] | 1 star | 1 star | 1 star | 1 star | 0 star | 1 star | 1 star | 0 star | 6 star |
| Butt, 2022 [73] | 1 star | 1 star | 1 star | 1 star | 1 star | 1 star | 1 star | 0 star | 7 star |
| Grewal, 2022 [101] | 1 star | 1 star | 1 star | 1 star | 1 star | 1 star | 1 star | 1 star | 8 star |
| Kissling, 2022 [122] | 1 star | 1 star | 1 star | 0 star | 1 star | 1 star | 1 star | 1 star | 7 star |

[a] Risk of bias assessment could not be performed for articles not published as full texts [65,71,75,81,90,97,125].

[b] 1 star was given if the study population was truly or somewhat representative of a community or population.

[c] 1 star was given if the nonexposed cohort was drawn from the same community as the exposed cohort. If the nonexposed cohort was drawn from a different source, no star was awarded.

[d] 1 star was given if a secure record (e.g., medical record) or a structured interview was used to ascertain exposure.

[e] 1 star was given if the outcomes were assessed at the beginning of the study.

[f] 1 star was given if the study was adjusted for age, sex, and marital status or other important factors. If the cohorts were not comparable on the basis of the design or the analysis did not control for confounders, no star was awarded.

[g] 1 star was given if the outcome was assessed by independent blind assessment or record linkage.

[h] 1 star was given if the duration of follow-up was sufficient.

[i] 1 star was given if there was complete follow-up or the lost to follow-up rate was ≤20%. No star was awarded if the follow-up rate was not reported.

[j] 1 star was given if the case definition was adequate and independently validated. No star was given if the case definition was based on record linkage, or self-reported, or not described.

[k] 1 star was given if the cases were consecutive or a representative series of cases. No star was given if there was a potential for selection biases or the representativeness of cases was not stated.

[l] 1 star was given if community controls were used. No star was awarded if hospital controls were used or controls were not described.

[m] 1 star was given if the control had no history of disease. No star was given if no description was provided.

[n] 1 star was given if the study controlled for the most important factor; 1 star was also given if the study controlled for any additional factors (up to 2 stars).

[o] 1 star was given if exposure was ascertained from a secure record or structured interview with blinding to the case/control status. No stars were given if this was ascertained by non-blinded interviews or written self-report or if no description was provided.

[p] 1 star was awarded if the method was the same. If a different method was used to ascertain cases and controls, no star was given.

[q] 1 star was given if the nonresponse rate was the same for both cases and controls. No star was given if the nonresponse rate was different or not described.

**Fig. S1** Summary of meta-analysis results on clinical effectiveness outcomes of the mRNA-1273 versus BNT162b2 COVID-19 vaccines in patients with the following cancers: hematologic malignancy, solid tumor, or mixed cancers.

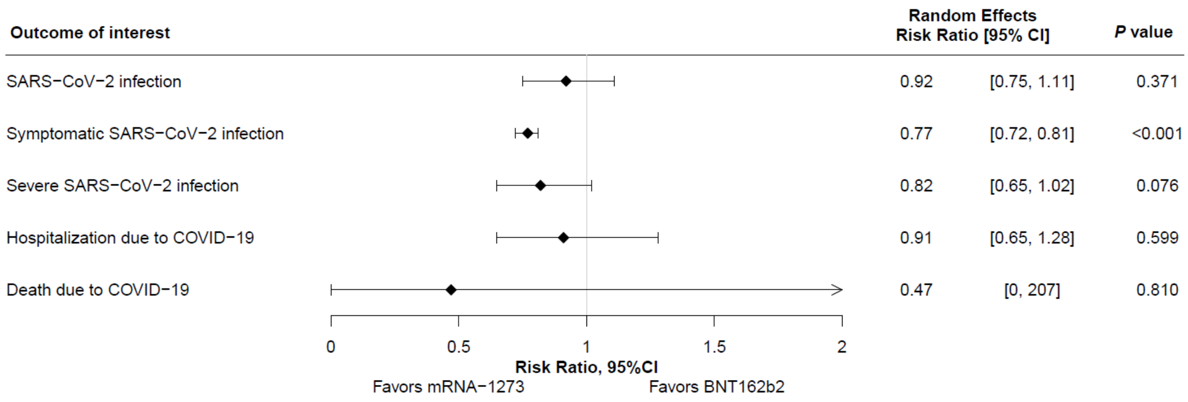

**Fig. S2** Summary of meta-analysis results on clinical effectiveness outcomes of the mRNA-1273 versus BNT162b2 COVID-19 vaccines in patients with the following cardiovascular-metabolic-renal diseases: cardiovascular disease, hypertension, type 1 or 2 diabetes, obesity, or chronic kidney disease with or without hemodialysis. NA, not applicable.

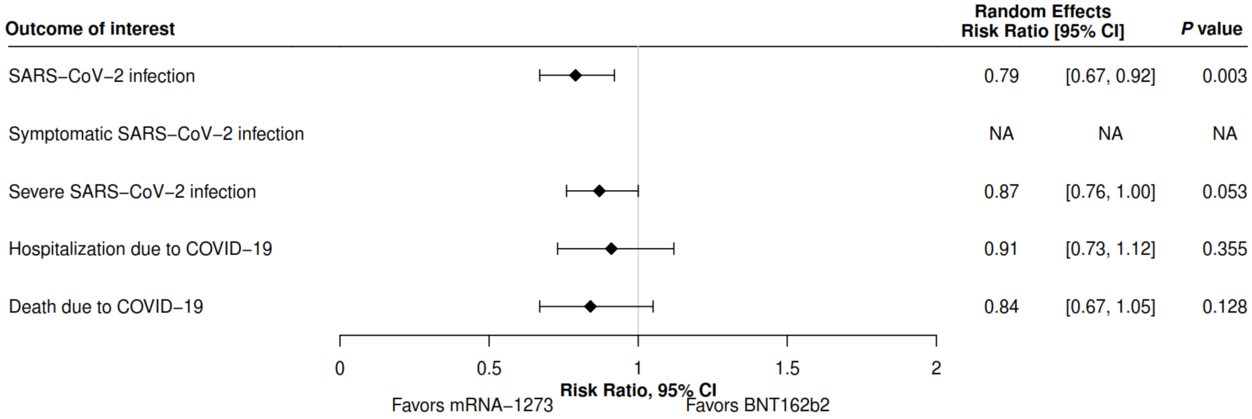

**Fig. S3** Summary of meta-analysis results on clinical effectiveness outcomes of the mRNA-1273 versus BNT162b2 COVID-19 vaccines in patients receiving the following immunosuppressive therapies: glucocorticoids, cytostatics, monoclonal antibodies, drugs acting on immunophilins, interferons, opiods, tumor necrosis factor-binding proteins, mycophenolate, or small biologic agents.

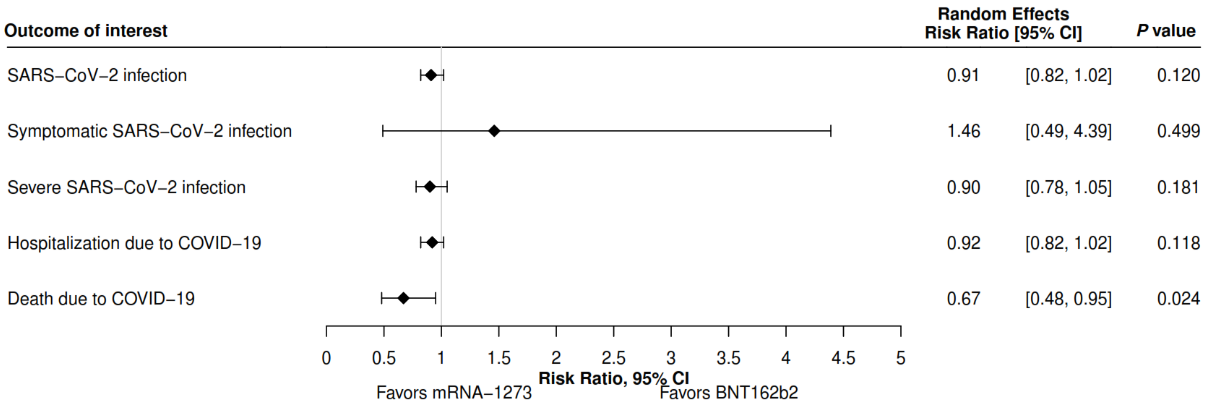

**Fig. S4** Summary of meta-analysis results on clinical effectiveness outcomes of the mRNA-1273 versus BNT162b2 COVID-19 vaccines in patients with the following autoimmune disorders: overall autoimmune diseases (rheumatic disease, dermatologic disease, neurologic condition); inflammatory bowel disease; inflammatory bowel disease (exposed to conventional and advanced immunosuppressive therapies); multiple sclerosis and on disease-modifying therapies; rheumatologic or inflammatory disorder; systemic lupus erythematosus; taking conventional synthetic DMARDs, biologic DMARDs, or glucocorticoids within 3 months of baseline; or systemic autoimmune rheumatic disease on immunomodulatory medications. DMARD, disease-modifying anti-rheumatic drug.

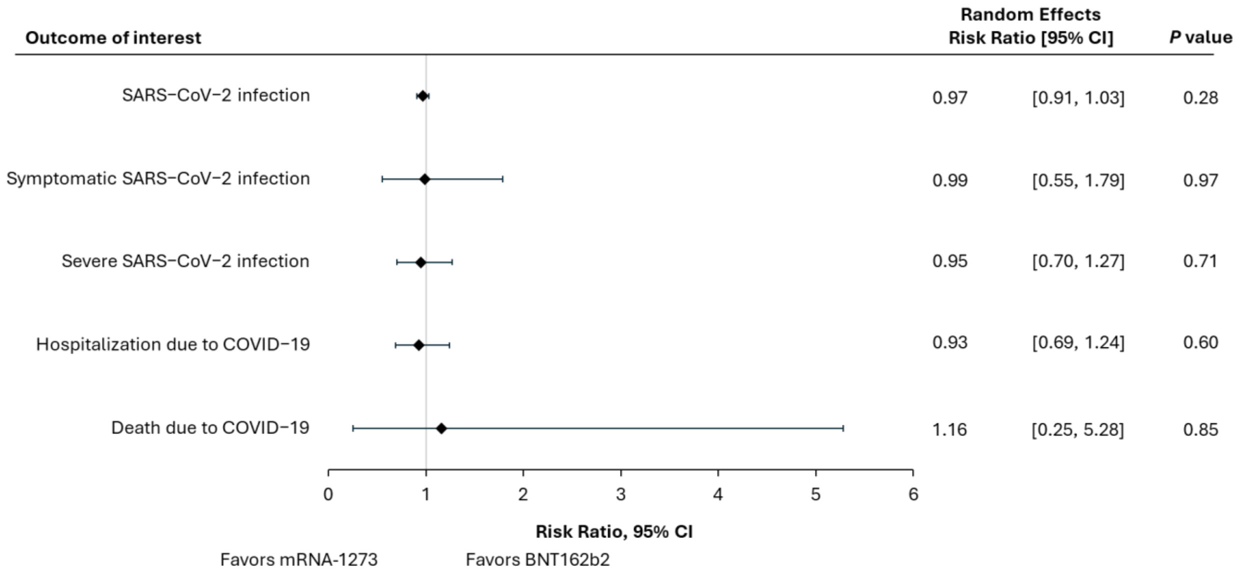

**Fig. S5** Summary of meta-analysis results on clinical effectiveness outcomes of the mRNA-1273 versus BNT162b2 COVID-19 vaccines in patients with cardiovascular disease (only hypertension). NA, not applicable.

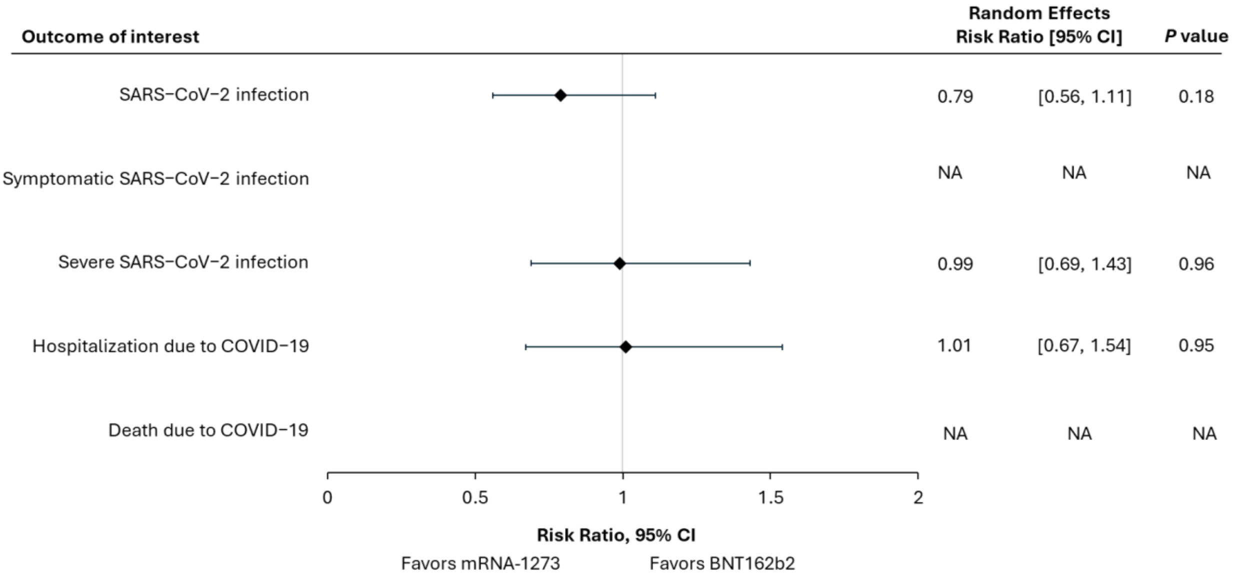

**Fig. S6** Summary of meta-analysis results on clinical effectiveness outcomes of the mRNA-1273 versus BNT162b2 COVID-19 vaccines in patients with chronic kidney disease. NA, not applicable.

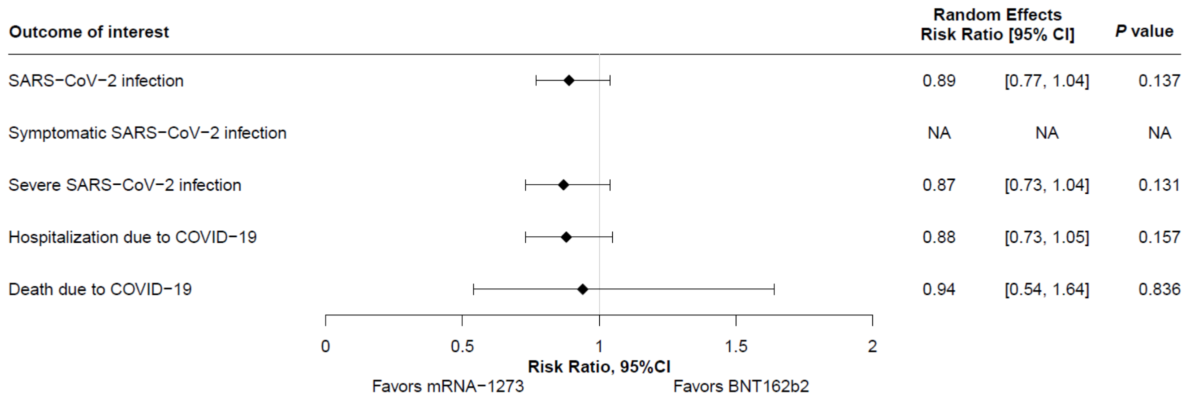

**Fig. S7** Summary of meta-analysis results on clinical effectiveness outcomes of the mRNA-1273 versus BNT162b2 COVID-19 vaccines in patients with chronic respiratory conditions. NA, not applicable.

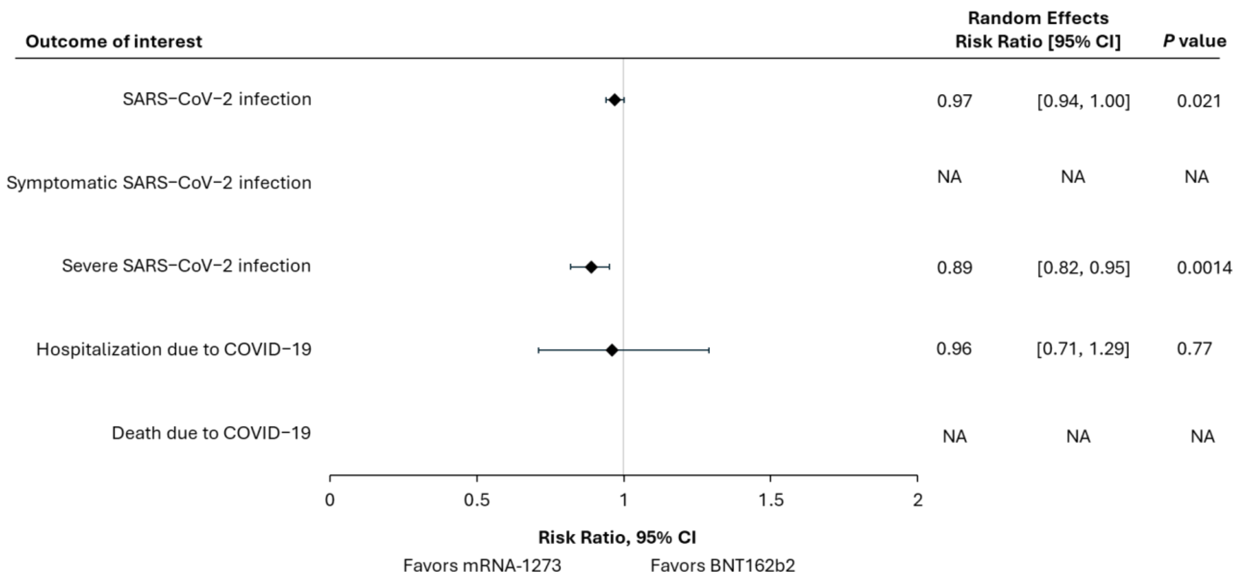

**Fig. S8** Summary of meta-analysis results on clinical effectiveness outcomes of the mRNA-1273 versus BNT162b2 COVID-19 vaccines in patients with diabetes. NA, not applicable.

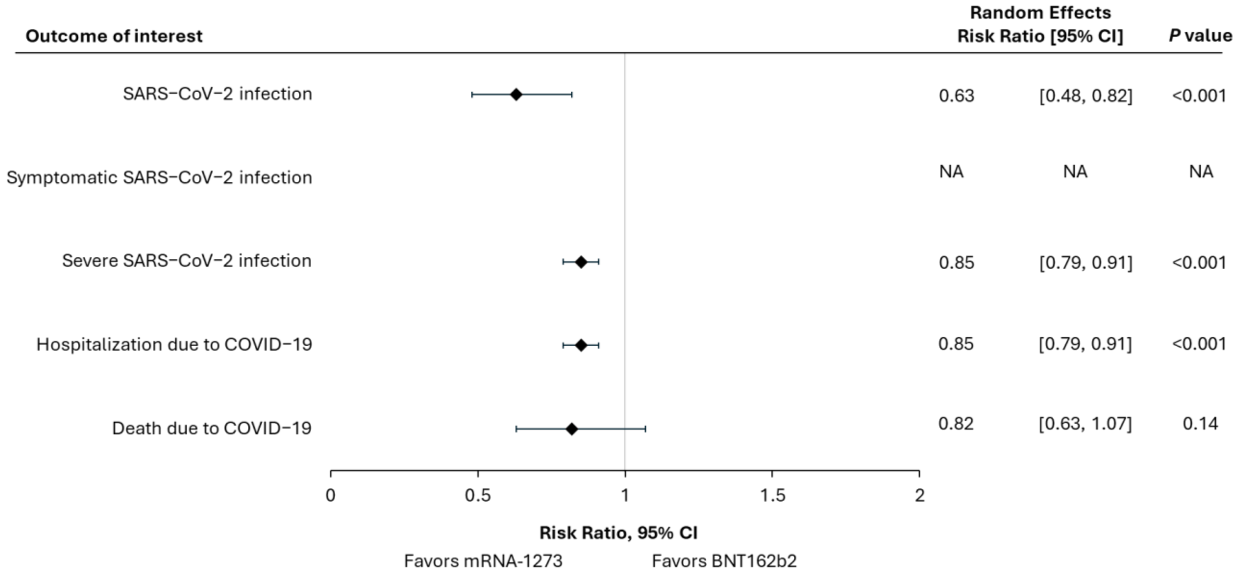

**Fig. S9** Summary of meta-analysis results on clinical effectiveness outcomes of the mRNA-1273 versus BNT162b2 COVID-19 vaccines in patients with hematologic malignancies. NA, not applicable.

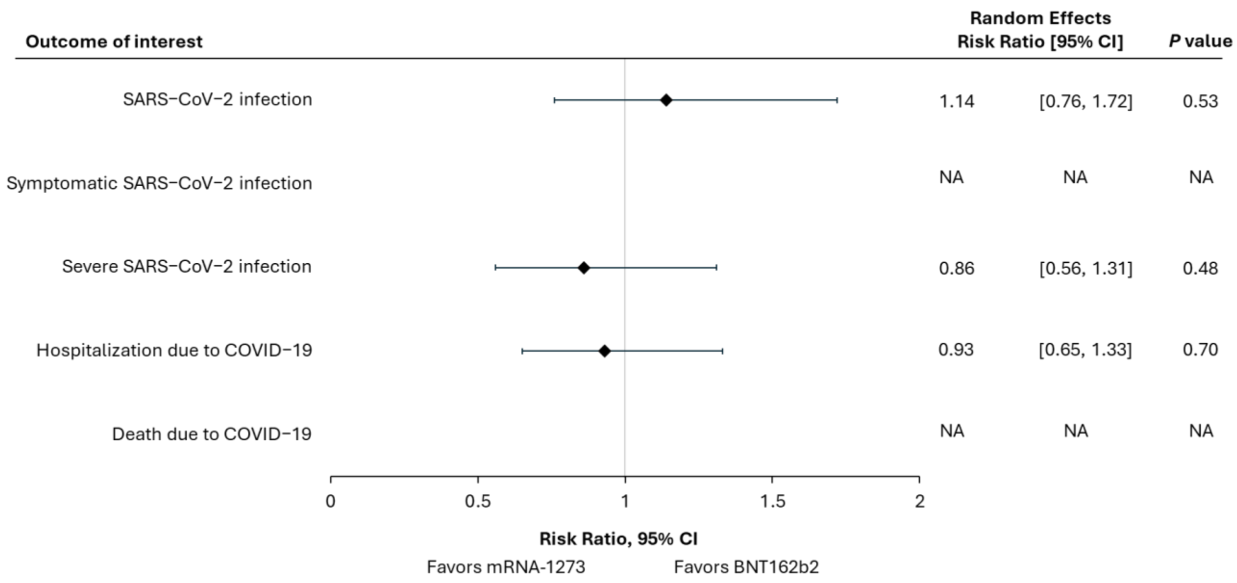

**Fig. S10** Summary of meta-analysis results on clinical effectiveness outcomes of the mRNA-1273 versus BNT162b2 COVID-19 vaccines in patients who received a solid organ transplant.

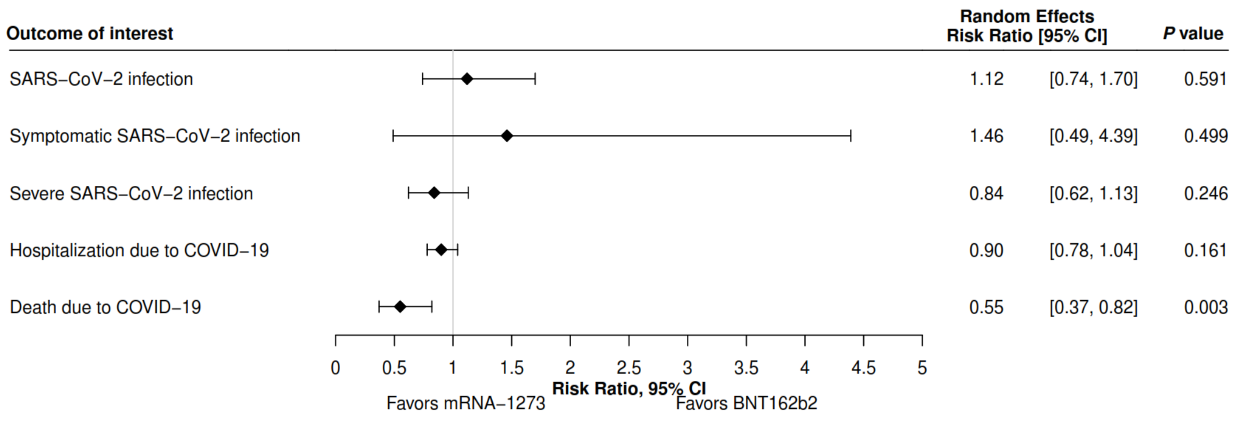
